# Evidence for Maternal Autoantibodies in the Pathogenesis of Preterm Birth

**DOI:** 10.1101/2024.10.03.24314850

**Authors:** Elze Rackaityte, Beltran Borges, Hannah M. Kortbawi, Haleigh S. Miller, Shirley J. Shao, Kelsey C. Zorn, Colette Caspar, Rebecca Wong, Frank McCarthy, Joseph S. Creery, Margareta Mayer, Elaine Kouame, Robert R. Puccinelli, Amy L. Kistler, David J.L. Yu, Sammer Roach Ganem, Alexandra DeRisi, Quinn Kawaja, Chung-Yu Wang, Aditi Saxena, Scott Oltman, Andrew F. Kung, Sabrina A. Mann, Anthea Mitchell, David Huang, Gabrielle Rizzuto, Rebecca J. Baer, Caleigh Mandel-Brehm, Kelli K. Ryckman, Stephanie L. Gaw, Sara L. Hillman, Laura L. Jelliffe-Pawlowski, Joanna Halkias, Nasim C. Sobhani, Mark S. Anderson, Francsesca Trespidi, Claudia Alpin, Tiziana Bosoni, Riccardo Albertini, Camilla Bellingeri, Barbara Gardella, Alessandro Borghesi, Nicole Prince, Jessica Lasky-Su, Michael R. Wilson, Marcelle Cedars, Joshua E. Elias, Tippi C. MacKenzie, Joseph L. DeRisi

## Abstract

Complications from preterm birth are the leading cause of global mortality in children under age five^1,2^. Spontaneous preterm labor is the most common cause of preterm delivery and is associated with a breakdown of maternal-fetal tolerance^3–5^. However, the current understanding of the role of autoantibodies in this process has been limited to a handful of examples of pathogenic antibodies that occur with pregnancy complications^6–14^. Here, we employ proteome-wide autoantibody profiling via phage display immunoprecipitation and sequencing (PhIP-seq) to identify autoreactivities associated with pregnancy as well as term or preterm delivery outcomes in maternal sera across eight cohorts of human pregnancy (n=2,194). We find that preterm pregnancies exhibit greater proteome-wide autoreactivity, validated by placental proteome immunoprecipitation mass spectrometry analysis using patient sera. Within the preterm birth associated autoreactive signature, we find enrichment for pathways known to be critical for normal pregnancy outcomes, including vascular development, reproductive hormones, and regulators of inflammation. Further analysis of autoreactive targets revealed involvement of the IL1β inflammatory cytokine cascade. IL1β is one of the few inflammatory cytokines sufficient to rapidly induce labor in animals^15–24^ and it is also elevated in preterm human pregnancies^25–29^. Across these eight cohorts, antibodies to cytokine IL1 receptor antagonist (IL1RA), a natural antagonist to IL1β are significantly enriched in roughly 7% of preterm pregnancies. Sera from these patients functionally neutralize IL1RA activity *in vitro* and anti-IL1RA induces greater resorption, inflammation, and vascular malperfusion in timed-pregnant mice *in vivo*. These findings suggest utility for serologic diagnostics as one approach to stratify risk of spontaneous preterm delivery in addition to interventional strategies for restoring control of IL1β during pregnancy.

## MAIN TEXT

Globally, 10% of pregnancies result in preterm delivery which corresponds to an annual 15 million preterm births^2^. Preterm newborns, born before 37 weeks, are disproportionately vulnerable to sepsis, neuroinflammation, and respiratory distress^2^. Despite the high mortality associated with these complications and the costly burden to healthcare systems, there are currently no FDA-approved medications to prevent preterm delivery available in the US market^30,31^. Tocolytic agents are used to transiently suppress preterm labor, and antibiotics may be administered in cases of preterm premature rupture of membranes (PPROM) to treat infection and indirectly reduce labor onset, but neither approach effectively prevents preterm birth. While several maternal^32^, fetal^33,34^, and environmental factors^35,36^ have been associated with preterm birth, the lack of mechanistic understanding of disease pathophysiology hinders the development of effective diagnostics and therapeutics.

Preterm birth is a common symptom of multiple discrete pathologies. While 30-35% of preterm births are medically induced for maternal or fetal indications^1^, the majority of preterm births (55-75%) occur spontaneously in the setting of preterm labor. These are also associated with infection and/or inflammation in approximately 30-40% of cases^1^.

Although little is known about the causes of preterm labor in this clinical subset, animal studies have demonstrated that certain inflammatory cytokines are sufficient to induce labor^20,37–39^. Intrauterine injection of inflammatory cytokines, particularly IL1β, rapidly induces labor in non-human primates^20–24^ and rodents^15–19^. This finding mirrors associations in humans, where elevated inflammatory cytokines are detected in mothers during labor, including spontaneous preterm labor^3,25–29^.

The failure to establish or maintain maternal-fetal immune tolerance is also associated with spontaneous preterm birth^40^. Recently, tolerization of maternal B cells through antigen masking on the placenta has been implicated in the establishment of maternal-fetal tolerance^41^. However, our current understanding of the humoral immune system in this process in humans has been limited to a handful of examples of pathogenic autoantibodies that occur with pregnancy complications, such as anti-RhD^6^, anti-thyroid^12^, anti-human platelet antigen^42^, and anti-AT1 receptor^13,14^ antibodies. Here, we describe the use of proteome-wide autoantibody profiling technologies to generate a comprehensive portrait of autoimmune reactivities in maternal sera that specifically precede preterm birth.

### Maternal autoantibody enrichment pregnancy atlas

To comprehensively investigate autoreactivity during pregnancy, maternal sera from term and preterm pregnancies derived from eight separate cohorts across three countries were used to determine antibody autoreactivity. The cohorts captured cross-sectional (I, IV, V), longitudinal (II, III, VI–VIII), and population-based (I) designs (**Fig 1a**). Cohorts III and VI comprised only term pregnancies with dense sampling across gestation. Paired maternal–cord blood samples were available for cohorts III and IV (**Extended Data Fig 1a-d; Supplementary Table 1**). Gestational age at sample collection was highly similar between preterm and term outcomes (**Fig 1b**, **Supplementary Table 1**; **Extended Data Fig 1a-b**). Nulliparous female and male samples from two cohorts of healthy individuals were used as additional controls (n=130). Serum from these cohorts was used to immunoprecipitate a customized, previously validated programmable phage display library of the human proteome with the PhIP-seq approach^43,44^. In total, 4,512 samples representing 2,324 individuals were analyzed.

**Figure 1.**
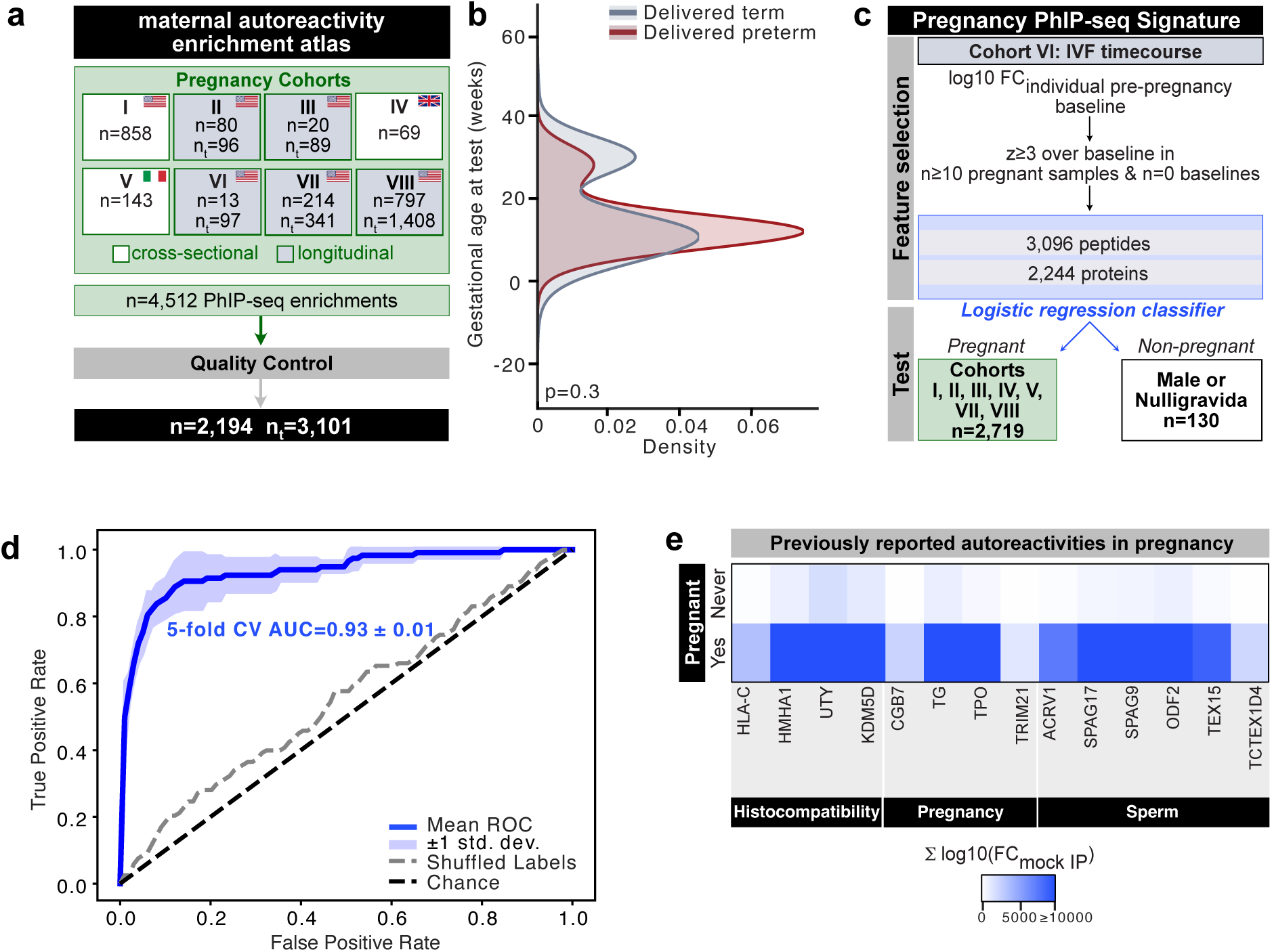
Maternal autoantibody signature induced by becoming pregnant. **a.** Schematic of study design integrating data from eight human pregnancy cohorts of maternal sera human analyzed by PhIP-seq with the human peptidome (n indicates human subject counts, n_t_ indicates human samples across multiple timepoints). **b**. Histogram of gestational age at time of sampling in weeks for women that delivered term and preterm across all cohorts tested. **c**. Schematic for identifying the PhIP-seq signature utilizing PhIP-seq data. Feature selection was performed on Cohort VI, where log10 fold changes were taken over individual pre-pregnancy baselines and z-scores were calculated over this baseline. Peptides were considered pregnancy-specific if they were detected in 10 pregnant samples and zero baselines. These features were used to train a five-fold cross validated logistic regression classifier to discriminate pregnant (Cohorts I-V,VII-VIII) and non-pregnant samples. **d**. Five-fold cross-validated performance of features selected from Cohort VI on pregnant (Cohorts I-V,VII-VIII) and non-pregnant samples. **e**. Sum fold change over mock immunoprecipitation of selected proteins in the PhIP-seq pregnancy signature in pregnant or never pregnant samples.Mann-Whitney U test for significance in b.

**Figure 2.**
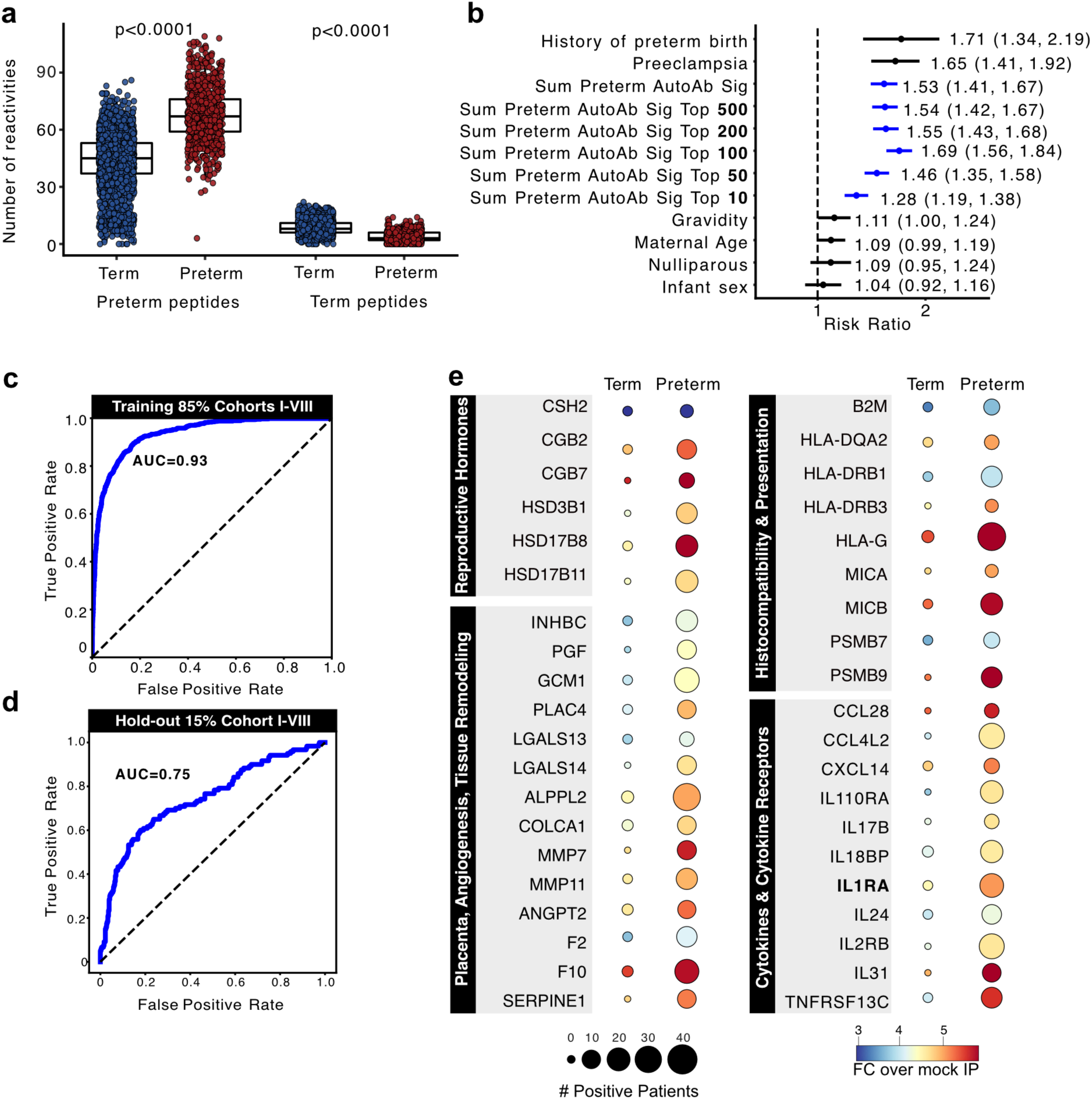
The preterm autoantibody signature. **a.** Number of preterm- and term-specific autoantibody reactivities per individual in term (blue) and preterm (red) pregnancies across all cohorts, **b.** Univariate forest plot of predictors for preterm birth including the preterm autoantibody signature. Odds ratios and 95% confidence intervals. Continuous predictors were scaled to one standard deviation; binary and categorical predictors were modeled relative to their reference group. The dashed vertical line indicates the null (OR = 1) on a logarithmic scale. Right labels report the point estimate and 95% Cl. Rows are sorted by effect size magnitude. Only samples for which all clinical metadata was available were included for this analysis. Receiver operator curve analysis of out-of-fold predictions from a forecasting model built on the preterm autoantibody signature in the **c** training set and **d** the hold-out set. **e.** Dot plot of selected autoreactive targets in healthy term (left) and preterm (right) pregnancies within selected pathways. Dots are scaled to the number of positive patients and colored by sum fold change above mock IP. PhlP-seq data filtered for conserved features across PhlP-seq was used to generate models and graphs. One-way ANOVA with Tukey HSD test for significance in a.

Following three rounds of phage enrichment, the encoded peptide inserts were sequenced and aligned to the input library to calculate overall enrichment as previously described^43^. A commercial anti-GFAP polyclonal antibody was used as a positive control across 96 well plates, batches, and cohorts (n=88). Peptides corresponding to the GFAP protein were enriched by an average of 10^3^-fold relative to IPs with sera from the human cohort samples (**Extended Data Fig 2a**). Prior to any filtering, PhIP-seq immunoprofiles from healthy controls were investigated for batch effects. A subset of samples (n=10) was technically replicated nine times in random positions across 20 experimental plates. Each set of ten technical replicates exhibited high Pearson co-correlation, and as has been previously observed for immune profiling of healthy controls^45^, comparison of individuals to each other yielded low to no correlation (**Extended Data Fig 2b**). After quality control filtering to remove under-sequenced (see Methods) and averaging technically replicated samples, a total of n=3,101 samples derived from n=2,194 pregnant subjects were retained.

### Global remodeling of autoreactivity during pregnancy

During pregnancy, the maternal immune system must adapt to accommodate the semi-allogeneic fetus. To first assess the induced proteome-wide autoreactivity that accompanies normal pregnancy, we analyzed a cohort of term pregnancies resulting from frozen embryo transfer (FET; Cohort VI) with sampling pre-FET, across gestation, and postpartum. Peptide-level fold changes were computed relative to each individual’s pre-pregnancy baseline. Pregnancy-specific peptides were defined as those with z-score ≥ 3, present in ≥ 10 pregnancy samples and absent from all baseline samples (**Fig 1c**). To derive the common signature of autoreactivity during normal pregnancy, the resulting 3,096 peptides, from 2,244 proteins, were used to build a logistic-regression classifier. The classifier was then used to distinguish pregnant (n=2,719) from non-pregnant (n=130) samples from all cohorts except Cohort VI, which was used during feature selection only. The mean AUC of the receiver-operating-characteristic analysis was 0.93 ± 0.01 by (ROC; **Fig 1d**), supporting the existence of a large set of shared autoreactive pregnancy-associated changes. The PhIP-seq pregnancy signature recapitulated previously reported reactivities to histocompatibility molecules, pregnancy-associated proteins, and sperm-antigens (**Fig 1e**) and identified autoreactive targets not previously known to be pregnancy associated (**Extended Data Fig 3, Supplemental Table 3**). Together, these data define a complex yet highly shared set of autoreactive changes in the pregnant mother.

### The preterm autoantibody signature

We next assessed whether preterm pregnancies exhibit a shared autoreactive signature distinguishable from term pregnancy. A set of healthy term pregnancies from Cohort I was selected controls (n=204; see Methods), and used to calculate mean RPK values at the peptide level.

Subsequently, peptide-level fold changes were calculated over the control means for all pregnancy cohorts. Peptides with a z-score of 3 or greater were designated as preterm-enriched if they were shared by at least four preterm subjects and were enriched in one or less of the 204 healthy term subjects (z-score ≤ 3; **Extended Data Fig 4**; see Methods). Conversely, term-specific peptides were identified using the mean of the preterm samples (down-sampled to n=204) to calculate z-scores (see Methods). This analysis yielded a total of 9,209 peptides derived from 3,594 proteins that were labeled as preterm-specific and 1,278 peptides derived from 868 proteins that were labeled as term-specific (**Supplementary Table 3**). The number of reactivities within the preterm autoantibody signature was significantly greater in preterm than term individuals. Conversely, the number of reactivities within the term autoantibody signature was overall lower, but still significantly higher in term than preterm pregnancies (**Fig 2a**). This difference in enrichments was consistent across all eight cohorts (**Extended Data Fig 5**). While the progression of normal pregnancy is associated with an increase in autoreactivity, the autoreactive enrichments derived from preterm individuals are both distinct and more numerous. The breadth of the preterm signature was also found to be significantly increased as a function of trimester at the time of sampling in preterm but not term pregnancies (**Extended Data Fig 6a-c**), with clear divergence already apparent in the first trimester.

We next assessed whether specific clinical features of preterm delivery were reflected within the preterm autoantibody signature. Summed enrichment of the preterm autoantibody signature was significantly greater in pregnancies with preterm deliveries before 28 weeks as compared to moderate or late preterm deliveries (**Extended Data Fig 7a**). Spontaneous preterm pregnancies exhibited a modestly higher preterm aut<u>o</u>antibody signature enrichment than iatrogenic preterm pregnancies (**Extended Data Fig 7b**). Beyond preterm birth subtypes, the preterm autoantibody signature was assessed for relationships with other pregnancy-related characteristics within preterm samples. Multigravidae, and especially women with 4 or more previous pregnancies, exhibited significantly higher autoreactivity than primigravidae (**Extended Data Fig 7c**), which was also true when considering an individual’s first pregnancy to those with one or more past pregnancies (**Extended Data Fig 7d**). C-section delivery, infant sex, history of preterm birth, and preeclampsia diagnosis did not significantly associate with the preterm autoantibody signature (**Extended Data 7e-h**). Thus, the preterm autoantibody signature is enriched in spontaneous and extremely early preterm births and increases with gravidity, but is otherwise independent of other measured clinical variables.

To determine whether the preterm autoantibody signature itself contributes to preterm birth risk relative to established obstetric risk factors, an odds ratio analysis was performed (**Fig 2b**). Each standard deviation increase in the summed log-fold change over mock IP was associated with significantly higher odds of preterm delivery (all features: OR = 1.53, 95% CI 1.41–1.67). To assess the strength of the association as a function of the number of autoantibody signature features, the feature set was restricted from as few as 10 peptides to as many as 500 peptides. Even with only 10 peptide features, the odds ratios were still significant (OR = 1.28, 95% CI 1.19–1.38), and by 100 features, the maximum odds ratio, comparable to the full autoantibody signature, was achieved (OR = OR = 1.69, 95% CI 1.56–1.84). Established obstetric risk factors showed the expected direction and magnitude: prior preterm birth (RR = 1.71, 95% CI 1.34–2.19) and preeclampsia (RR = 1.65, 95% CI 1.41–1.92) were each associated with increased risk. In contrast, nulliparity (RR = 1.09, 95% CI 0.95–1.24), infant sex (RR = 1.04, 95% CI 0.92–1.16), maternal age per standard deviation (OR = 1.09, 95% CI 0.99–1.19), and gravidity per standard deviation (OR = 1.11, 95% CI 1.00–1.24) were not clearly related to preterm birth risk. Altogether, the composite autoantibody signature yields an effect size comparable in magnitude to major clinical risk factors, with performance improving as more signature peptides are included.

To orthogonally assess the presence of antibody-bound placental proteins, a randomly selected subset of sera from Cohort II (n=13 term, n=25 preterm) was used to immunoprecipitate native proteins from placental extract. Antibody targets were identified using trapped ion mobility time of flight mass spectrometry (timsTOF-MS; see Methods). Mass spectrometry-derived protein abundances were used to calculate z-scores relative to term pregnancies (see Methods). A total of 343 preterm proteins were identified by IP-MS (**Extended Data Fig 8a-b**; see Methods). While PhIPseq displays linear peptides, IP-MS may also capture full-length proteins and conformational epitopes. Despite this difference, 65% (n=224 proteins) were also found by PhIP-seq in this cohort (**Extended Data Fig 8b**; **Supplementary Table 4**). A UMAP analysis of the enriched protein abundances yielded two distinct clusters, distinctly segregating preterm or term samples (**Extended Data Fig 8c**). Top ranked proteins by prevalence revealed sharing across multiple individuals, and hierarchical clustering of these features clearly separated preterm versus term sera (**Extended Data Fig 8d**). While neither PhIPseq or IP-MS distinguishes between maternal or fetal derived antigens, the combined PhIPseq and IP-MS data both support the notion that auto- and allo-reactive antibodies describe preterm pregnancy.

### Training and validation of a predictive preterm birth model

The preterm autoantibody signature suggested that we could develop a model predictive of impending preterm birth using a serological assay. As a proof of concept, a logistic regression model was constructed to predict preterm delivery from PhIP-seq profiles. Data were partitioned into sets for feature selection and model training (n = 2,655), with representative samples from each cohort held out for independent validation (n = 475; **Extended Data Fig. 9a–b**). During the feature selection phase, we performed 1,000 random five-fold cross-validation splits using the training set. Within each fold, autoreactive peptides were selected if they exceeded the z-score thresholds as above (see Methods). Gestational age at sampling was included as a co-variate to account for gestation-dependence in autoreactive profiles across cohorts (**Extended Data Fig 6**). Features were evaluated for stability across training iterations. Across 1,000 model iterations, 889 features were stable at the 80% threshold, 2,000 features at 50%, and 7,337 features at 25%, with only three peptides appearing in every iteration (**Extended Data Fig 9d**). These stability-selected features were then used to train a new five-fold cross-validated model, which yielded out-of-fold ROC AUC values of 0.933 for the 25% stability set, 0.914 for the 50% stability set, 0.797 for the 80% stability set, and 0.583 for the global stability set (**Fig 2c**, **Extended Data Fig 9e**). Given the best training-set performance of the 25% stability feature set, this model was selected.

To assess whether predictive performance was influenced by clinical or demographic variables, we stratified model accuracy across key subgroups. The model discriminated spontaneous preterm deliveries with slightly higher accuracy (ROC AUC = 0.88) than iatrogenic preterm deliveries (ROC AUC = 0.84; **Extended Data Fig 9f**). Limited change in performance was observed in samples collected across trimesters of pregnancy and cord blood (ROC AUC range =0.76-0.79; **Extended Data Fig 9g**). These analyses indicate that the classifier captures an autoreactive signature of preterm birth that is largely independent of etiology or demographic variables.

The final trained model was utilized to predict preterm or term delivery outcomes of the hold-out set (n=475), which was never used for training or feature selection. As expected, model performance on the validation cohorts was less than the training cohorts, however, strong forecasting ability was still achieved (ROC AUC 0.75; **Fig 2c**; **Supplementary Table 5**). The analysis of model performance also suggests that the immunological changes associated with preterm birth are varied and confirm the necessity for numerous and diverse cohorts for training.

### Targets within the preterm autoantibody signature

The features that constitute the preterm autoantibody signature correspond to a rich landscape of proteins distributed across structures and functions related to pregnancy and the fetus. Identification of most prevalent features (**Supplementary Table 3**) associated with risk included PLA2G2A, a secreted phospholipase involved in prostaglandin biosynthesis, a process that is essential for placental adaptation and labor onset among other roles and IZUMO3, classically a sperm protein^46^, raising the possibility of alloantibody responses to paternal antigens.

Pathway enrichment analysis was also used to aggregate and summarize relationships between preterm autoantibody signature features (Methods). These pathways included hormone synthesis and signaling, placental angiogenesis and tissue remodeling, histocompatibility, and cytokine categories (**Fig 2e, Extended Data Fig 10**). Several critical reproductive hormones (and/or their receptors) were significantly enriched (CSH2, CGB2, CGB7, PGF). Vascular remodeling of maternal spiral arterioles is a hallmark of successful placentation^47^. Accordingly, proteases (e.g. MMP11, MMP7) and canonical angiogenesis proteins (e.g. LAMB3) were significantly targeted in preterm pregnancies (**Fig 2e**).

Preterm autoantibody targets in the infection response category also included histocompatibility and antigen processing proteins (B2M, HLA-DPB1, HLA-DQA2, MICA, MICB, PSMB9) and the placental-specific HLA-G (**Fig 2e** **Supplementary Table 6**). Anti-HLA antibodies increase the risk of spontaneous preterm delivery^48^ and are associated with complement deposition in the placental endothelium, which occurs more frequently in spontaneous preterm labor^7^. Anti-cytokine antibodies, known to increase susceptibility to infection^49^, were also identified. Of these, anti-IL1RA and anti-IL1R2 were the most significantly enriched. Furthermore, anti-IL1RA was the only anti-cytokine reactivity that was detected both by PhIP-seq and IP-MS (**Fig 2e**; **Suplementary Table 3; Supplementary Table 4**). Together, the essential roles of these proteins, their expression on placenta, and their enrichment by preterm sera suggests that beyond a breakdown in tolerance, there is the possibility that some of the corresponding autoantibodies may possess pathologic function-blocking activities related to pathways that maintain successful pregnancy.

### Neutralizing anti-IL1RA autoantibodies define a subset of preterm pregnancies

Our comprehensive profiling of autoreactivity during pregnancy uncovered a heterogeneous immune landscape, with preterm pregnancies exhibiting patterns that distinctly diverge from those seen in term pregnancies. To better understand autoreactivities that may contribute to the pathogenesis of preterm birth, we screened for proteins with established roles in the initiation of parturition and their corresponding antagonists. We purposely included low-prevalence hits, especially those targeting secreted proteins. Among candidates, we identified interleukin-1 receptor antagonist (IL1RA; encoded by the IL1RN gene) as a possible lead. IL1RA is the secreted competitive antagonist of IL1β, whose T labor-promoting effects are well-established^20^; the existence of autoantibodies to the secreted competitive antagonist, IL1RA, has been noted in other contexts^50–53^. Despite low prevalence by PhIPseq across all cohorts (0.3%; **Fig 3a**), we pursued studies of anti-IL1RA as an exemplar of preterm autoreactivity with a plausible mechanistic consequence.

**Figure 3.**
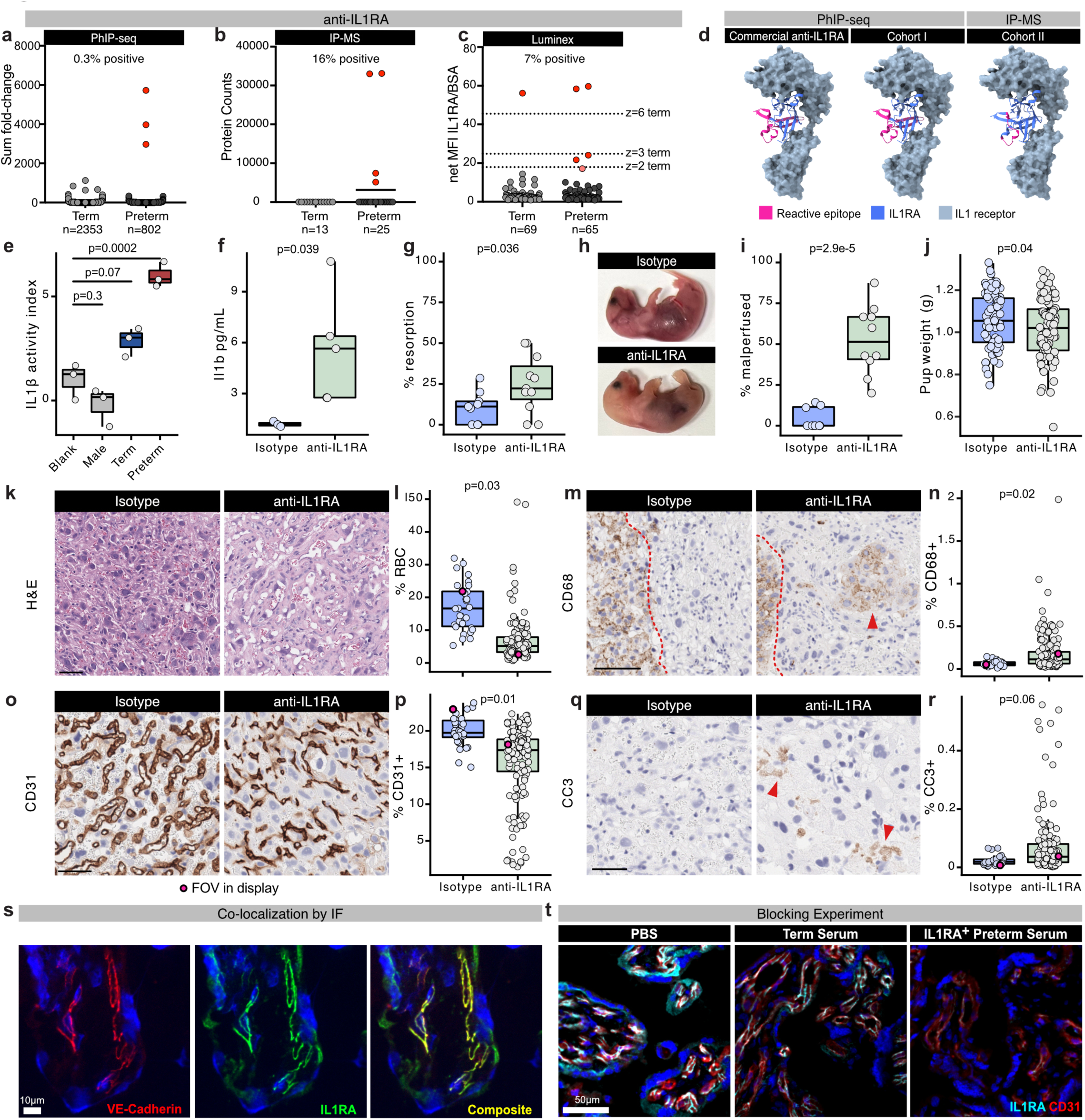
Anti-IL1 RA autoantibodies in pregnancy. Anti-IL1 RA antibodies in preterm versus term pregnancies identified using a. fold change above mean in term pregnancies detected by PhlP-seq, b. protein counts detected by placental IP-MS in Cohort II, or c. net MFI of IL1RAover BSA detected by Luminex assay in Cohort V. **d.** Location of reactive epitope (magenta) determined by PhlP-seq orIP-MS for an anti-IL1 RA commercial antibody or enriched in preterm sera in Cohort I or Cohort II on the structure ofhuman IL1 RA (green) bound to the IL1 receptor (blue). PDB accession: 1IRA. e. Quantification of secreted embryonic alkaline phosphatase (SEAP) produced by HEK-Blue IL-1 β reporter cells incubated with human IL-1 β and IL-1RAin the presence of male, term, anti-IL1RA^+^ sera or no serum controls. IL1 activity was calculated by dividing experimental signal with patient sera by cells treated with IL1 alone, **f.** Quantification off. 111 b (pg/mL) in murine maternal serum by ELISA, **g.** complete resorption rate (%), h. representative images of non-resorbed pups, i. percentage of malperfused pups among live pups in isotype or anti-lL1 RA injected dams at E18.5 supplemented with 5µg of IL1 β. Micrographs of placentas stained with **k.** H&E, **m.** anti-CD68, **o.** anti-CD31, and **q.** anti-cleaved caspase 3 (CC3) from isotype or anti-IL1RA injected dams at E18.5 supplemented with 5µg of IL1 β Quantification of **I.** red blood cell (RBC), **n.** CD68+, **p.** CD31 +, **r.** CC3+ area in each field of view (FOV); magenta dot denotes field of view in display. Spinning disk confocal micrographs of indirect immunofluorescence of placenta probed with s. anti-VE-cadherin (left) or anti-l L1 RA antibody (middle). Composite (right) indicates overlay of signal from anti-VE-cadherin and anti-IL1RA. **t.** Spinning disk confocal micrographs of indirect immunofluorescence of placenta preblocked with PBS, term sera, or anti-IL1RA^+^ preterm sera from Cohort II. Overlay of signal from anti-CD31 (red), anti-IL 1 RA (cyan), and DAPI (blue). DAPI counterstain in blue; scale bar corresponds to 10µm in k,m,o,q and 5Oµm in t. Statistical significance calculated by one-way ANOVA with Tukey post-hoc in e or Kolmogorov-Smirnov pairwise test in f,g,i. FOV quantifications were averaged and per-mouse Kolmogorov-Smirnov pairwise test is reported in I, n, p, r.

To assess the prevalence of anti-IL1RA antibodies, two orthogonal approaches were used. First, immunoprecipitation-mass spectrometry (IP-MS) was used to interrogate Cohort II sample (20–30 weeks gestation; n=25 preterm, n=13 term). This analysis recovered antibody bound IL1RA in 20% of samples (**Fig 3b**). In an independent cohort (Cohort V; 11–15 weeks gestation; n=65 preterm, n=69 term), we implemented a Luminex assay which detected anti-IL1RA in 7% of preterm pregnancies and <1.5% of term pregnancies (**Fig 3c**). We further profiled 894 longitudinal samples from Cohort VIII (n=86 preterm, n=808 term) using the same Luminex platform. Prevalence in preterm pregnancies increased from 3.3% at 10–12 weeks to 8.0% at 12–14 weeks’ gestation. In contrast, term pregnancies exhibited <2% prevalence before 15 weeks gestation. We also noted an increase in anti-IL1RA positivity among term pregnancies at 30 weeks (6.7%) (**Extended Data Fig 11a-b**).

To assess whether IL-1RA autoantibody positivity was associated with timing of delivery, we fit a logistic regression model. Among antibody-positive individuals, earlier gestational age at first positivity trended toward higher odds of preterm birth (OR per week earlier=1.6, 95 % CI 0.85–3.1, p = 0.14). In a model including never-positive pregnancies, the risk of pre-term delivery increases as the inverse of gestational age at which anti-IL1RA is first detected (OR = 2.12, 95% CI 1.89-2.38, p < 10-35; **Extended Data Fig 11c-d**). Together, these results support a model in which early emergence, rather than presence alone, is associated with increased preterm risk.

To identify the specific epitope on IL1RA that is targeted by anti-IL1RA antibodies, peptide fragments from PhIP-seq and IP-MS were analyzed. Positional mean enrichment analysis of PhIP-seq data (see Methods) revealed that the majority of anti-IL1RA^+^ pregnancies targeted overlapping peptides between D99-D129 amino acid residues on IL1RA. This epitope (**Fig 3d**; **Extended Data Fig 12a**), opposite the receptor-engaging face, is highly charged and contains the only N-linked glycosylation residue in the protein (N109)^54,55^. Antibodies to this epitope have previously been reported in IgG4 disease^51^ and it is also targeted by two commercial antibodies (**Fig 3d**; **Extended Data 12a**). IP-MS fragment analysis identified a single 16 amino acid peptide (103-LQLEAVNITDLSENRK-118) that was fully contained within the target PhIP-seq fragment (**Fig 3d**; **Extended Data 12a**). These data confirm the target epitope enriched by both PhIP-seq and IP-MS are localized to the same region, and furthermore, these data suggest that anti-IL1RA antibodies present in multiple pre-term sera converge on the same region of the protein.

To investigate the functional consequences of anti-IL1RA antibodies, a previously described IL1β reporter cell line and validated assay was utilized^50–52^. Briefly, neutralization of IL1RA was measured using a reporter cell line in which IL1 receptor activity results in secreted embryonic alkaline phosphatase (SEAP). Reporter cells were stimulated with recombinant IL1β and a titration of increasing amounts of excess of IL1RA and serum or antibody (**Extended Data Fig 12b-d**), SEAP rates were normalized to the negative control (TNFα treated cells) and an IL1β activity index was calculated for cells stimulated with IL1RA and IL1β relative to IL1β alone (see Methods). Compared to cells treated with PBS or off-target anti-GFAP antibody, a commercial anti-IL1RA antibody (R&D AF-280-NA) that primarily targets the patient epitope (**Fig 3d**), resulted in significant neutralization of IL1RA as measured by greater IL1β activity (**Extended Data Fig 12c-e**). Similarly, anti-IL1RA^+^ preterm sera also resulted in significantly higher IL1β activity (**Fig 3e**; **Extended Data Fig 12d**).

Conversely, cells treated with PBS, male, or term sera exhibited low IL1β activity. Thus, both commercial antibodies and preterm patient sera are sufficient to neutralize IL1RA antagonist, thereby permitting increased IL1β signaling.

### Maternal anti-IL1RA antibodies promote fetal resorption and malperfusion in vivo

To further explore the physiological consequence of circulating anti-IL1RA antibodies *in vivo,* the role of maternal anti-IL1RA antibodies was investigated during murine pregnancy, in which exposure to IL1β results in reproductive defects^19^. We hypothesized that since IL1RA regulates IL1β, adoptive transfer of anti-IL1RA antibodies would inhibit the ability of IL1RA to rescue pregnancies threatened by IL1β-induced inflammation, resulting in worse pregnancy outcomes. First, we confirmed that the patient epitope targeting commercial anti-IL1RA antibody was cross-reactive to the native mouse orthologue by dot blot (**Extended Data Fig 13a**). Timed-pregnant dams were injected intravenously three times (Embryonic Days 13.5-15.5; E13.5-15.5) with either the anti-IL1RA antibody or an equivalent amount of isotype control in the context of IL1β-induced inflammation (**Extended Data 13b**). Levels of murine il1β were measured in maternal serum by ELISA on the day before expected delivery (E18.5). Concentration of maternal Il1β was significantly greater in anti-IL1RA injected dams as compared to isotype controls (**Fig 3f**), indicating that anti-IL1RA neutralizes the ability of the antagonist to regulate Il1β during pregnancy. Consistent with our hypothesis, anti-IL1RA injected dams also exhibited profound reproductive defects. Resorption rate was significantly elevated (**Fig 3g**). Among the non-resorbed pups, a significantly greater proportion of fetuses from anti-IL1RA pregnancies exhibited less prominent cephalic vasculature and pallid skin color (**Fig 3h-i**; **Extended Data Fig 13c**), suggestive of a defect in placental perfusion. Non-resorbed pups from anti-IL1RA pregnancies weighed significantly less than isotype-injected controls (**Fig 3j**). Malperfused pups weighed significantly less than normal pups, though a defect in gross placental weight was not observed (**Extended Data 13d-f**). Histological analysis of placenta revealed reduced red blood cell perfusion into the placental labyrinth in animals treated with anti-IL1RA (**Fig 3k**). Placentas from anti-IL1RA treated dams also exhibited vessel constriction (lower CD31+ area), immune infiltration (greater CD68+ cells), and elevated apoptosis (cells with cleaved caspase 3), as determined by immunohistochemistry (**Fig 3m-r**).

Anti-IL1RA impaired IL-1 signaling and produced pregnancy pathology even without exogenous IL1β, but both factors contributed to resorption phenotypes. IL1β supplementation had a significant effect (p = 0.008), and its interaction with anti-IL1RA was also significant (p = 0.04; **Extended Data Fig. 13g**). Maternal IL1β levels were driven by both IL1β dose and antibody administration (**Extended Data Fig. 13h**). In contrast, fetal malperfusion was highly specific to anti-IL1RA, as IL1β dose, alone or in interaction, did not significantly explain variance (**Extended Data Fig. 13i**). These patterns extended to fetal and placental weights: placental weights were reduced by IL1β, whereas fetal growth restriction tracked with malperfusion (**Extended Data Fig. 13d–f**). Together, these results show that anti-IL1RA is sufficient to disrupt IL-1β regulation and induce vascular and fetal defects, but pathology is most severe when IL1β is concurrently dysregulated.

### IL1RA is expressed in adherens junctions of placental vessels

To investigate localization of IL1RA in human placenta, we used indirect immunofluorescence with a commercial antibody to IL1RA (Sigma Prestige HPA001482). A striking pattern of IL1RA localization was found near CD31-positive fetal vessels within the placenta (**Extended Data Fig 13a-c**) that also co-localized with VE-cadherin in endothelial cell junctions (**Fig 3s**). While IL1RA is known to be secreted, these results suggest a substantial amount of this protein is localized to the endothelial adherens junctions in the placenta.

To address whether the observed pattern of IL1RA co-localization was authentic, or due to cross reactivity of the antibody with other targets (such as VE-cadherin), PhIP-seq was performed with the same antibody and analyzed for enrichment. Only the IL1RA derived peptides were enriched above background, suggesting the commercial IL1RA antibody is highly specific (**Extended Data Fig 13d**). As a further validation of antibody specificity, a blocking assay was performed with the peptide immunogen used to generate the antibody (**Extended Data Fig 13e**). The signal corresponding to IL1RA was eliminated, again suggesting this antibody is highly specific (**Extended Data Fig 13f**).

Together, these data support the notion that IL1RA is bound at placental endothelial cell junctions, in close proximity to other junction proteins, like VE-cadherin.

To determine whether anti-IL1RA antibodies functionally bind placenta, we investigated whether anti-IL1RA^+^ patient sera were capable of blocking IL1RA on placental tissue from binding by a commercial antibody. A positive result would further confirm the existence and specificity of anti-IL1RA antibodies with respect to native placental tissue-bound IL1RA protein. Flash frozen placenta was first blocked with either PBS or term or anti-IL1RA^+^ patient sera, and then indirect immunofluorescence was performed with the commercial anti-IL1RA antibody (see Methods; **Extended Data Fig 13g**). The characteristic pattern of IL1RA localization to fetal vessel cell-cell junctions was clearly observed in the absence of sera, term sera or anti-IL1RA^-^ preterm sera (**Fig 3t**; **Extended Data Fig 13g**).

However, IL1RA localization signal was blocked in placental tissue treated with anti-IL1RA^+^ preterm sera (**Fig 3t**; **Extended Data Fig 13g**). These data strongly support the presence of highly specific anti-IL1RA^+^ autoantibodies in the maternal sera of a subset of preterm pregnancies, and that these antibodies readily bind native tissue-bound IL1RA in normal placenta.

## DISCUSSION

Certain pathogenic alloantibodies are known to drive pregnancy complications in humans^6,12,13^, motivating a systematic examination of maternal autoreactivity during gestation. By profiling more than two thousand maternal sera across eight pregnancy cohorts, we defined a comprehensive landscape of humoral autoreactivity in term and preterm pregnancies. Overall autoreactivity increased with advancing gestational age and gravidity, underscoring the dynamic and evolving nature of the maternal immune repertoire. After adjusting for gestational age at sampling, we identified a preterm birth-associated autoantibody signature that was consistently enriched across geographically and ethnically diverse cohorts. Notably, these reactivities were detected an average of fifteen weeks before preterm delivery, indicating that serologic changes precede clinical onset rather than result from it. Because preterm birth arises from diverse pathophysiological causes, including inflammatory, vascular, and idiopathic etiologies, the associated autoantibody landscape is correspondingly heterogeneous, likely contributing to the large feature set required for classification. The autoreactivies that drive the classification model may be mechanistically pathogenic and/or reflect broader immune dysregulation.

One limitation of this work is that the model does not reveal the underlying triggers or events that lead to defects in self-tolerance associated with preterm birth. However, within this broader signature, multiple proteins comprise plausible autoreactive targets of functional significance. Among these, a rare subset of preterm pregnancies harbored functionally inhibitory antibodies against the IL-1 receptor antagonist (IL1RA) that exacerbated IL-1β–driven reproductive defects in mice and were capable of binding IL1RA in human placenta. Their presence may reflect post-infectious or autoimmune immune priming, as anti-IL1RA antibodies have been described after COVID infection^52^, and in IgG4^51^ and Still’s disease^53^. Together, these findings support a two-hit model, in which pre-existing or infection-induced dysregulation of IL-1β signaling is amplified by anti-IL1RA antibodies, tipping the balance toward inflammation, placental injury, and pregnancy loss.

Although the pathogenic role of these antibodies requires further investigation, recombinant IL1RA (anakinra; SOBI, Inc.) has been used clinically for over two decades with established long-term safety in non-pregnant populations^56,57^ and limited case reports of use in pregnancy^58^. These data suggest that anakinra use in pregnancy to prevent preterm labor merits further evaluation, particularly in stratified cohorts defined by anti-IL1RA antibody status.

Beyond an individual protein, this collective proteome-wide data, representing thousands of individuals, are a rich compendium of human humoral autoreactivity in normal and preterm pregnancy, making it ideal for emerging machine learning approaches, especially with respect to classification tasks. As human antibody and antigenic repertoire techniques continue to also evolve, it is likely that clinically useful descriptors of immune dysregulation will also emerge, as will targeted therapeutic interventions to limit the effects of autoantibodies, which in turn may yield improved management of pregnancies at risk for preterm delivery and reductions in neonatal morbidity and mortality.

## METHODS

### Human cohorts and samples

Sera from the Cohort I was obtained from a California-wide biobank (Committee for the Protection of Human Subjects within the Health and Human Services Agency of the State of California protocol# 12-09-0702). Sera from the Cohort II, Cohort III, Cohort VI, and Cohort VII (UCSF IRB# 10-00505, 20-31171, 20-32077, 20-32779, 16-20474; San Francisco, CA) was obtained from deliveries at UCSF. Sera from Cohort IV (UCSF IRB# 10-00350) was obtained from UCL Hospital and Homerton University Hospital of the National Health Services of the United Kingdom. Sera from Cohort V was obtained from ‘Fondazione IRCCS Policlinico San Matteo’ (San Matteo Research Hospital) in Pavia, Italy, under RC08061819 and RC08061821 approved IRB protocols. Sera from Cohort VIII (IRB# 2009P000557 and 2014P001109, Boston, MA) was obtained from Brigham and Women’s Hospital in Boston, MA, USA from the previously described VDAART clinical trial^59^. De-identified healthy control (non-pregnant) plasma was collected from two sources: courtesy of New York Blood Center (New York, NY) and donors from a UCSF community drive (UCSF IRB# 22-3611; San Francisco, CA). All samples were collected under the referenced Institutional Review Board protocols, and all patients gave informed consent prior to sample collection.

Uncomplicated, term pregnancies were selected from Cohort I to define a baseline of autoreactivity. An uncomplicated, term pregnancy for this cohort was defined as one resulting in delivery at greater than or equal to 37 weeks of gestation and did not have a diagnosis of any of the following: placental abruption, placenta previa, chorioamnionitis, oligohydraminos, polyhydraminos, premature rupture of membranes, bacterial vaginosis, urinary tract infection, diabetes or gestational diabetes, preexisting or gestational hypertension, preeclampsia, infection during pregnancy, any specified placental condition, retained placenta with or without hemorrhage, unspecified hemorrhage, abnormal clotting, or threatened abortion. Any pregnancy that required transfusion, rhesus isoimmunization, or cerclage was excluded from this group. We also excluded mothers with any hypertension disorder, endometriosis, Group B streptococcus positivity, any coagulation deficiency, malignancy, asthma, allergic dermatitis, anaphylaxis, rheumatoid arthritis, systemic lupus erythematosus, autoimmune thyroiditis, or mental disorder. Mothers who reported any smoking, drug or alcohol abuse or dependency during pregnancy were excluded. Pregnancies where the infant exhibited convulsions, abnormal neural imaging or exam, retinopathy of prematurity, respiratory distress syndrome, intraventricular hemorrhage, necrotizing enterocolitis, bronchopulmonary dysplasia, periventricular leukomalacia, hypoxic-ischemic encephalopathy, or known congenital heart disease were also excluded from this group. Finally, we also required that women defined as having uncomplicated pregnancies did not have a history of recurrent pregnancy loss, preterm delivery, or poor obstetric/reproductive outcomes.

Term pregnancies for all cohorts were defined as deliveries at 37 or greater weeks of gestation.

Preterm pregnancies were defined as those delivering prior to 37 weeks of gestation and were further subdivided into spontaneous or iatrogenic preterm delivery. Spontaneous preterm pregnancies were defined as pregnancies with spontaneous labor with delivery prior to 37 weeks of gestation. Iatrogenic preterm pregnancies were defined as pregnancies with delivery prior to 37 weeks of gestation and without spontaneous labor.

### PhIP-seq with human peptidome library

The human T7 phage display library used for immunoprecipitation and sequencing is previously described and sera from the above cohorts was used in high-thruput protocols as previously described. Detailed protocols are published on protocols.io DOI: dx.doi.org/10.17504/protocols.io.4r3l229qxl1y/v1

### Trapped Ion Mobility Time of Flight Mass Spectrometry of Immunoprecipitated Placental Protein Lysate

Term placenta from a single donor was harvested within 2 hours of delivery. Implantation side of placenta was dissected to 1 cm^3^ blocks and flash frozen in liquid nitrogen. Placenta protein lysate was prepared by mechanically dissociating tissue using a glass Dounce homogenizer and an electric tissue homogenizer in RIPA buffer with protease inhibitors (Roche) on ice. Protein was quantified using Bradford Assay (Pierce), normalized to 500µg in TNP40 and incubated with 1µL of human sera overnight at 4C with overhead mixing. Proteins were subsequently incubated with a 1:1 mix of protein A and protein G Dynabeads (Thermo) for 1 hour and washed five times with RIPA and once with TrisHCl. A NanoDrop reading was taken under TrisHCl to quantify protein.

Supernatant was removed and proteins were then resuspended in 8M urea, 50mM Tris (pH 8) before being subject to a standard on-bead protein digestion. The protein digestion procedure included disulfide reduction with 5 mM DTT, alkylation with 14 mM iodoacetamide, and an overnight digestion with Lys/C at a protein to protease ratio of 1:50. In order to obtain an optimal tryptic digestion, the urea concentration was then diluted to 2 M with 50 mM Tris (pH 8) followed by the addition of trypsin to a protein to protease ratio of 1:50, and a 4 hour incubation at room temperature. The peptide supernatant pH was then decreased to 2 with 7 µL formic acid (FA) and separated from the beads with a magnet. An extraction was performed on the beads by resuspension with 0.1% FA, the supernatant of which was added to the final sample. Peptide desalting was performed with the AssayMAP Bravo (Agilent Technologies) using a standard RPS peptide cleanup cartridges and protocol^60^. Approximately 25 ng of desalted peptides were analyzed on a TIMSTOF SCP mass spectrometer (Bruker Corporation) coupled with an EASY-nLC 1200 LC system (Thermo Fisher Scientific). Peptides were separated by reverse-phase chromatography on a 25 cm column (75-μm inner diameter, packed with 1.6 μm C18 resin, AUR2-25075C18A-CSI; IonOpticks). Peptides were introduced into the mass spectrometer using a gradient starting with 2% to 8% buffer B (0.1% (v/v) formic acid in 80% acetonitrile) for 1 min followed by an increase to 25% buffer B for 25 min then an increase to 40% buffer B for 5 min at a flow rate of 100 nL/min and were ionized by CSI (captive spray ionization).

Samples were first analyzed by PASEF-dda with a duty cycle of 1.03 sec comprising one MS1 survey for every 5 PASEF MS2 ramps and a mass range between 100 and 1700 m/z. The TIMS device ramp/accumulation time was set to 166 ms with a 100% duty cycle and a mobility range of 0.7 to 1.3 1/K0. Precursor ions from the survey scan were selected using an adjusted isolation width of 2 m/z below 700 m/z and 3 m/z above 800 m/z precursor mass-to-charge value in the quadrupole that aligns with the anticipated TIMS elution time relative to the ion mobility value. Fragmentation was performed by Collision Induced Dissociation (CID) with CE (collisional energy) interpolation between 42 eV at 0.65 1/K0, 31.92 eV at 0.8 1/K0, 36.96 eV at 1 1/K0, 42 eV at 1.2 1/K0, 47.04 eV at 1.4 1/K0 and 51.24 eV at 1.6 1/K0. Active exclusion of precursors were released after 0.2 minutes. Precursor repetitions were set to a target intensity of 20000 and a threshold of 500.

Real-time searching was performed with PaSER (Parallel Search Engine in Real-Time) against a human Uniprot database (downloaded on 30 July 2022) using a reverse-decoy method with default settings. A spectral library was generated from the searched DDA data using PaSER. Samples were then analyzed by PASEF-dia using a high speed acquisition scheme from Meier F et al and searched in real-time against the DDA generated spectral library using the TIMS DIA-NN algorithm with default settings.

Peptide counts were collapsed to the corresponding protein, log-normalized, and fold changes and z-scores were calculated over mean signal in term pregnancies. Preterm-specific hits were identified by requiring that at least one preterm pregnancy and no term pregnancies met the threshold of z>=5 for a given protein.

### Placental Immunofluorescence

Placental tissue was harvested within 2 hours of delivery and embedded, unfixed, in O.C.T compound (Tissue-Tek) and flash frozen in an isopentanol bath submerged in liquid nitrogen. Ten-micron cryosections were cut from frozen blocks and adhered to glass slides. Slides were thawed and briefly post-fixed with acetone, rehydrated in 1x PBS, blocked with 10% (v/v) goat serum in 1x PBS for 2 hours, and incubated with 1:1000 rabbit anti-human anti-IL1RA (Millipore Sigma Cat No. HPA001482) and 1:500 rat anti-human CD31 (Abcam Cat. No. ab9498) overnight at 4C in a humidified chamber.

Primary antibodies were washed using standard protocols and incubated with anti-rabbit and anti-rat secondaries using standard protocols. Secondary only controls were stained identically but omitting primary antibody. Stained tissue was imaged using the Crest LFOV Spinning Disk/ C2 Confocal at 400x magnification under identical camera exposure and laser settings for secondary only and experimental samples. Micrograph exposure was normalized to secondary only negative control in FIJI and applied to all images at once. In peptide blocking experiments, IL1RA PrEST Antigen (Millipore Sigma APREST83081) was incubated with anti-IL1RA antibody per manufacturer’s instructions at 4C overnight and used to stain tissue as above. In serum blocking experiments, placental sections were blocked with 1% (v/v) goat serum and 10% human serum in PBS-T overnight at 4C. Controls were blocked without human serum. Placenta was subsequently probed with the commercial anti-IL1RA antibody (Millipore Sigma Cat No. HPA001482) and imaged as above.

### In vitro IL1RA blocking assay

Diluted patient sera, anti-IL1RA (R&D AF-280-NA), or anti-GFAP were pre-incubated with human IL1-RA (Peprotech) overnight and applied to the HEK-Blue IL1β cells (InvivoGen) for 2 hours. Cells were subsequently stimulated with human IL1β (Peprotech) for 72 hours at 37C. Cells treated with IL1β alone or TNFa alone (Peprotech) were used as positive and negative controls, respectively.

Supernatants were assayed using the QANTI-Blue assay (InvivoGen), as previously described^51,52^. IL1β activity index was calculated by measuring SEAP activity at OD_655_ every 15 minutes for 2 hours and determining the rate of enzymatic activity. Slopes were background subtracted (cell treated with TNFa) and normalized to mean signal in IL1β treated cells. IL1β activity index was defined as the fold change of signal in serum or antibody treated cells over signal in cells incubated with IL1RA and IL1β only. Each experiment was carried out in triplicate for a total of three biological replicates.

### Murine Il1ra dot blot

500 or 250 nanograms of recombinant mouse Il1ra (R&D 480-RM-050) were blotted onto nitrocellulose using a custom 3D printed dot blot device under vacuum. Membranes were blocked for 1 hour at room temperature in 5% (w/v) milk and subsequently probed overnight at 4C with either anti-human IL1RA antibodies (R&D AF-280-NA or Sigma Prestige HPA001482) or anti-mouse Il1ra antibodies (Invitrogen PA5-21776 or R&D MAB4801) or no primary antibody. Following standard washing procedures, membranes were probed with relevant secondaries conjugated to Licor-IR dyes and blots were imaged using the Licor Odyssey machine. Membranes were imaged together in one scan under identical exposure settings.

### Animal Husbandry and Injections

All mice were housed, bred, and maintained in a pathogen-free facility at the University of California San Francisco (UCSF). All procedures were performed in concordance with UCSF Institutional Animal Care and Use Committee (IACUC) regulations and approved protocol. Timed-pregnant C57BL/6 females mated with C57BL/6 males were purchased from Jackson Laboratories (strain #000664). Retro-orbital (RO) injections were done following with UCSF IACUC procedural guidelines. Briefly, pregnant dams were anesthetized at E13.5-E15.5 with isoflurane and injected in the RO sinus using 0.5mL insulin syringes, with a maximum volume of 150µL containing 0 or 5µg of human IL1β (Peprotech 200-01B) and 50µg of polyclonal goat IgG (isotype control; R&D AB-108-C) or of goat anti-human IL1RA antibody (R&D AF-280-NA) in sterile PBS. Reagents were mixed immediately prior to injection.

### Mouse harvesting

Mice were euthanized at E18.5 with CO_2_ and a sample of serum was obtained via transcardial puncture. Resorbed and malpefrused fetuses were quantified. Whole fetuses and their placentas were harvested and weighed. Murine Il1b concentrations were measured in maternal serum by a commercial ELISA (Abcam Cat No. ab197742).

### Histological analysis of murine placenta

Whole placentas were fixed in 4% paraformaldehyde and embedded in paraffin using standard protocols. Embedding, grossing, sectioning, immunohistochemistry (IHC) with anti-human antibodies to anti-CD68, anti-cleaved Caspase-3, or anti-CD31, and scanning was performed at HistoWiz.

Blinded H&E slides were assessed by a licensed pathologist. IHC was quantified by defining 5 non-overlapping regions in each of the two placental sections for a total of 10 regions for each placenta in QuPath software. Regions were randomly drawn and covered most of the placental area, avoiding anomalous regions (e.g. small tears in tissue). For anti-CD68 IHC, regions of interest were drawn in the labyrinth region of the placenta. A pixel thresholding classifier was built for each of the antibody stains using a Laplacian of Gaussian prefilter setting. DAB positive area was calculated as a percentage of total area within the defined region. Each individual region of interest was reported for isotype or anti-IL1RA treated conditions.

### Luminex assay to detect anti-IL1RA antibodies in patient sera

Recombinant IL1RA (Peprotech) or BSA (Thermo) were conjugated to spectrally-distinct Luminex beads in separate 1.5mL protein LoBind tubes. Each bead conjugation was performed as previously described^62^ using the Antibody Coupling Kit following manufacturer’s instructions (Luminex, 40-50016). All serological analyses were performed exactly as previously described^62^ in technical duplicate on two separate experimental days. Positive samples (with greater than 20 net MFI IL1RA/BSA) were repeated on an additional experimental day in technical duplicate. All net MFI IL1RA/BSA values were averaged and z-scores above mean in term were calculated.

### Bioinformatic Analyses

To analyze peptide enrichment after PhIP-seq, reads were aligned at the protein level using RAPsearch^63^, as previously described . Aligned reads were normalized to 100,000 reads per k-mer (RPK) to account for varying read-depth. All downstream analyses were performed by using an implementation of PhagePy python package (https://github.com/h-s-miller/phagepy).

All data was initially filtered to samples with fewer than recovered 100,000 reads. Data was pseudocounted and fold change over mock IP controls was calculated. Co-correlations of the fold change over mock IP matrix were used to determine technical replicate consistency. Subsequently, technically replicated samples (with the exception of positive and mock IP controls) were averaged.

Fold change over mock IP was re-calculated with averaged values. Additional fold changes were calculated over mean in healthy term pregnancies from Cohort I only (see above), preterm pregnancies (downsampled to n=204, five times, then averaged), or over all term pregnancies. Z-scores were calculated for each of the fold-change matrixes after log-10 transformation relative to mock IP, healthy term pregnancies, preterm pregnancies, and over all term pregnancies. Peptides with a z-score greater than 3 in a minimum of 4 experimental samples and 0 control samples were considered specific for that comparison, unless otherwise indicated. Analysis was performed on all cohorts together, except when performing comparisons for Cohort II PhIP-seq data with Cohort II IP-MS data, where the same analysis was performed using samples only from Cohort II. Autoreactivities were counted per person by summing up binarized per peptide hits within the preterm- or term-specific PhIP-seq signature. Where indicated, cumulative enrichment was calculated by summing log-transformed enrichments over mock IPs for peptides identified as preterm- or term-specific.

Positional enrichment analysis across IL1RA was performed as previously described^65^ for Cohort I samples. GSEA analysis was performed by manually annotating genes with previously reported roles in placental function and pregnancy. GO term enrichment analysis supplemented manual grouping and was performed utilizing the gseapy package.

### Machine Learning Predictive Modelling

#### Data and preprocessing

Gestational age at sampling was normalized to the average term pregnancy (40 weeks for all samples) and included as a co-variate. Clinical outcome labels were encoded as binary preterm birth (ptb: 1 = preterm, 0 = term). One auxiliary grouping variable was constructed to support fold-change estimation and stratified splitting: hc_term (1 = healthy term controls from Cohort I; 0 = all preterm).

#### Fold-change matrices

Peptide-level fold change matrices were computed as above over healthy term controls. Unless otherwise noted, downstream predictive modeling used this log-transformed matrix as features, and a sparse “selection-only” matrix of binarized peptides with a z-score greater than 3 over healthy term controls to drive in-fold feature selection (see below).

#### Train/holdout partitioning

Samples were split once into train and holdout sets. Splits were stratified by outcome and constrained by cohort and gestational-age bins ( bins = 12, 16, 20, 24, 28, 32, 36, 40 weeks). When available for longitudinal cohorts, subject barcode column was used to prevent leakage by keeping all samples from a subject within a single partition. All model development and cross-validation used only the training partition; final performance was assessed on the untouched holdout.

#### Feature construction and covariates

For modeling, the feature design matrix was built log10-transformed fold change over healthy term controls and augmented with the prespecified normalized gestational age at time of sampling, which was always included. The feature selection matrix was binarized peptides with a z-score greater than 3 over healthy term.

#### In-fold supervised feature selection (dual-matrix)

We implemented a fast, deterministic, in-fold selection that evaluates binary peptide “hits” from the selection matrix while training models on the continuous log10-transformed fold change matrix. Prior to cross-validation, the selection matrix was binarized once (hit_cutoff = 0.0).

Within each training fold, features were retained if they met:

*minimum hit count in preterm = 4, and*

*maximum hit count in term (any term sample) = 12.*

If no features met criteria, a fallback chose the top fallback_k = 50 features ranked by (case_hits − control_hits). A hard cap of max_features_per_fold = 5000 limited the per-fold dimensionality. Selection yielded names that were mapped to column indices of the FC matrix; GA_test_norm was appended post-selection. All selection occurred inside each training fold to avoid information leakage. The selection logic was encapsulated in a scikit-learn–compatible transformer (DualMatrixFoldSelector).

#### Classifier and hyperparameters

The core classifier was logistic regression with L2 penalty (solver="lbfgs", C=1.0, max_iter=2000) and class_weight="balanced". Features were imputed with a constant fill (0.0) and standardized (z-score) within each fold. All preprocessing steps (selection → imputation → scaling → classifier) were wrapped in a scikit-learn Pipeline to ensure proper cross-validation hygiene.

#### Cross-validation, out-of-fold (OOF) predictions, and model selection

To perform feature selection, we ran 1,000 outer iterations of 5-fold stratified CV on the training set (StratifiedKFold with shuffling; per-iteration seed i + 3). For each iteration and fold:

The fold-specific feature set was selected using the training portion only.

The pipeline was fitted and used to produce OOF probabilities for the held-out fold.

Across iterations, we recorded:

per-fold ROC-AUC and PR-AUC,

per-iteration mean ROC-AUC (averaged over the 5 folds), and the full OOF probability matrix (iterations × samples).

We also retained the per-fold selected feature lists and summarized their stability (see below). For qualitative reference, we saved the best and worst single fold models by ROC-AUC across all iterations.

#### Feature stability, intersections, and frequency panels

From all selection outputs (iteration × fold), we computed:

i. the global intersection (features present in every fold of every iteration),
ii. per-iteration intersections (features present in all 5 folds of an iteration), and
iii. frequency of selection across all folds. We defined stability panels at thresholds of ≥80%, ≥50%, and ≥25% of folds. We also constructed a frequency-top-N panel (e.g., top 500 by fold count). All panels included normalized gestational age at sampling at training time.

#### Panel training, coefficients, and OOF performance

For each panel, we trained a fixed-column logistic-regression pipeline (column locking → imputation → scaling → LR) and generated 5-fold OOF predictions on the training set (one repeat). We reported OOF ROC-AUC and PR-AUC and plotted OOF ROC curves. Final panel models were then refit on all training samples and saved with their locked column order. We exported standardized coefficients (per-SD) and approximate raw-scale coefficients together with the intercept to facilitate interpretation.

#### Holdout evaluation and subgroup analyses

Each saved panel model was evaluated on the held-out test set created by Step1_split. We computed ROC-AUC and rendered holdout ROC curves. To examine performance heterogeneity, we generated stratified ROCs contrasting preterm subtypes versus term (PTB type: iatrogenic, spontaneous) and timepoint (First, Second, Third, Cord_blood). For subgroup plots, we applied the same prediction vectors but stratified truth labels and sample indices per category. For all ROC curves, the trained model generated a predicted probability of preterm birth for every sample, and the ROC-AUC was calculated directly from these predicted probabilities against the true binary outcome.

#### Reproducibility and implementation details

All analyses were performed in Python using AnnData for matrix management and scikit-learn for model building. Randomness entered only through the CV shuffling seeds and the initial train/holdout split; fixed seeds are stated above. No global prefiltering was applied to the FC feature space beyond per-fold selection.

#### Reporting

All reported cross-validated metrics are computed strictly within the training partition using OOF predictions to avoid optimistic bias. Holdout metrics are computed once on the untouched test set. Subgroup ROCs are descriptive and based on the same prediction vectors, stratified by the indicated metadata fields.

### Data Availability

All raw and processed data, the forecasting model and feature weights, as well as the associated code are available for download at Dryad. Link for peer review: http://datadryad.org/stash/share/gOsb-l9_flhFstHmz1-CuX7BpqmGOf1DF1qQEX-UVK0. PhIP-seq analytical code is freely available on GitHub: https://github.com/h-s-miller/phagepy

## Data Availability

All data produced in the present study are available upon reasonable request to the authors

## ACKNOWLEDGEMENTS

We thank members of the DeRisi Lab for helpful discussions and JM Rackaitis for support during these studies. We thank Joelle Ostroff for inspiring the studies herein. We also acknowledge the New York Blood Center for contribution of healthy control plasma. We thank the UCSF CALM-NIC microscopy core for use of the CREST LFOV Spinning Disk/C2 confocal funded by the UCSF Program for Breakthrough Biomedical Research, the Sandler Foundation, Strategic Advisory Committee, and the EVCP Office Research Resource Program Institutional Matching Instrumentation Award. ER is funded by 2022 Next Gen Pregnancy Initiative Research Grant from the Burroughs Wellcome Fund, the Eunice Kennedy Shriver National Institute of Child Health and Human Development award 5T32HD098057, and National Institute of Allergy and Infectious Diseases award K99AI182451. CMB is funded by The Emiko Terasaki Foundation (project 7027742/fund B73335) and by the National Institute of Neurological Disorders and Stroke of the NIH award K99NS117800. GR is funded by National Institute of Allergy and Infectious Diseases award K08AI137209 and 2023 Next Gen Pregnancy Initiative Research Grant from the Burroughs Wellcome Fund. HK is supported by NICHD F30HD117526. SLG is supported by National Institute of Allergy and Infectious Diseases award K08AI141728 and R01HD111582. The Host-pathogen group (AB) at Fondazione IRCCS Policlinico San Matteo, Pavia, Italy, is supported by grants from the Italian Ministry of Health, RC08061819 and RC08061822, and by 5X1000 grant 08061821from the San Matteo Hospital. NP and JLS are supported by National Heart Lung and Blood Institute award R01HL169300. JLD is supported through funding from the Chan Zuckerberg Biohub. Contents herein are the sole responsibility of the authors and do not necessarily represent the official views of the NIH or other funding agencies.

## AUTHOR CONTRIBUTIONS

ER and JLD designed the research. ER, BB, HMK, HSM, SJS, KCZ, RW, FM, JSC, MM, EK, RP, AK, DJLY, CC, SG, AD, QK, GW, AS, SAM, AM, GR performed the research. ER, HMK, HSM, AFK, CMB, JSC, FM, JEE, and JLD contributed to new reagents and analytic tools. SO, RW, DH, RJB, KKR, SLG, SLH, LLJP, JH, NCS, FT, CA, TB, RA, CB, BG, AB, NP, JLS, MC, provided clinical samples, metadata, and/or contributed to clinical interpretation of data. ER, JLD, TCM, MSA, MRW provided supervision of the work. JSC, FM, and JEE generated and analyzed mass spectrometry datasets. ER, BB, and TCM provided significant contributions to mouse model development. ER and JLD analyzed data and produced figures. ER and JLD wrote the manuscript. All authors reviewed the manuscript and provided feedback.

## MATERIALS AND CORRESPONDENCE

Correspondence and requests for materials should be addressed to Joseph L. DeRisi.

