## Extended Data figures for "Evidence for Maternal Autoantibodies in the Pathogenesis of Preterm Birth"

#### Extended Data Figure 1

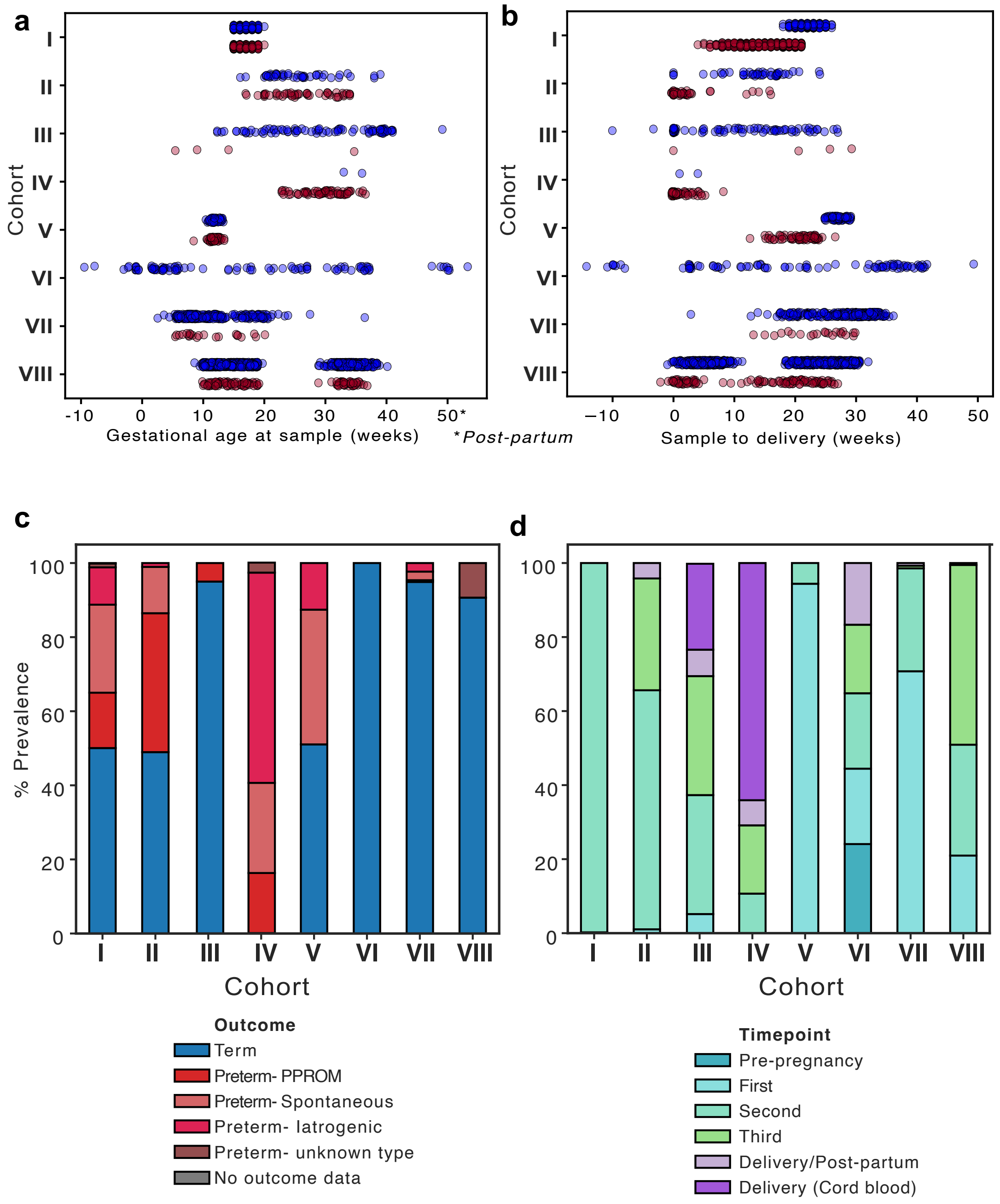


**Extended Data Figure 1**. **Maternal serum samples used in the study** Dot plot of gestational age **a.** at test or **b.** sample to delivery in weeks across the eight pregnancy cohorts with term (blue) and preterm (red) outcomes. Stacked barplots of **c.** pregnancy outcomes and **d**. timepoints at sample collection for eight pregnancy cohorts. Individual human samples represent each dot in a and b.

#### Extended Data Figure 2

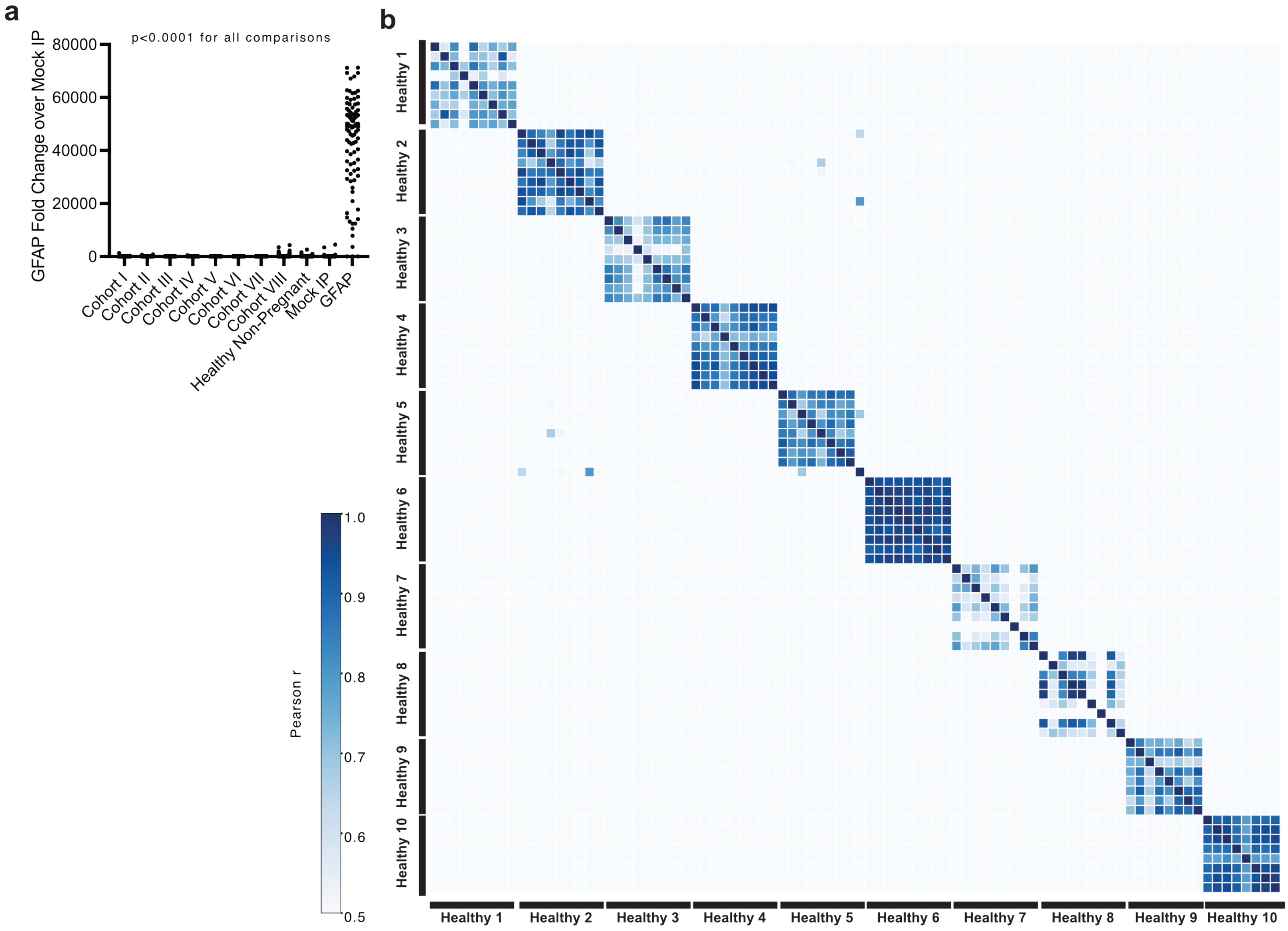


**Extended Data Figure 2**. **Technical consistency of the PhIP-seq method. a.** Sum reads per 100,000 (RPK) of peptides tiling across the GFAP protein in immunoprecipitations (IPs) with sera from Cohorts I-VII, healthy non-pregnant samples, mock IP, and anti-GFAP antibody. **b.** Pearson co-correlation analysis of fold change over mock IP across 10 technically replicated samples across 20 plates. Enrichments identified by PhIP-seq in a-b. Each dot represents an independent IP in a. ANOVA test for significance with Tukey post-hoc in a.

#### Extended Data Figure 3

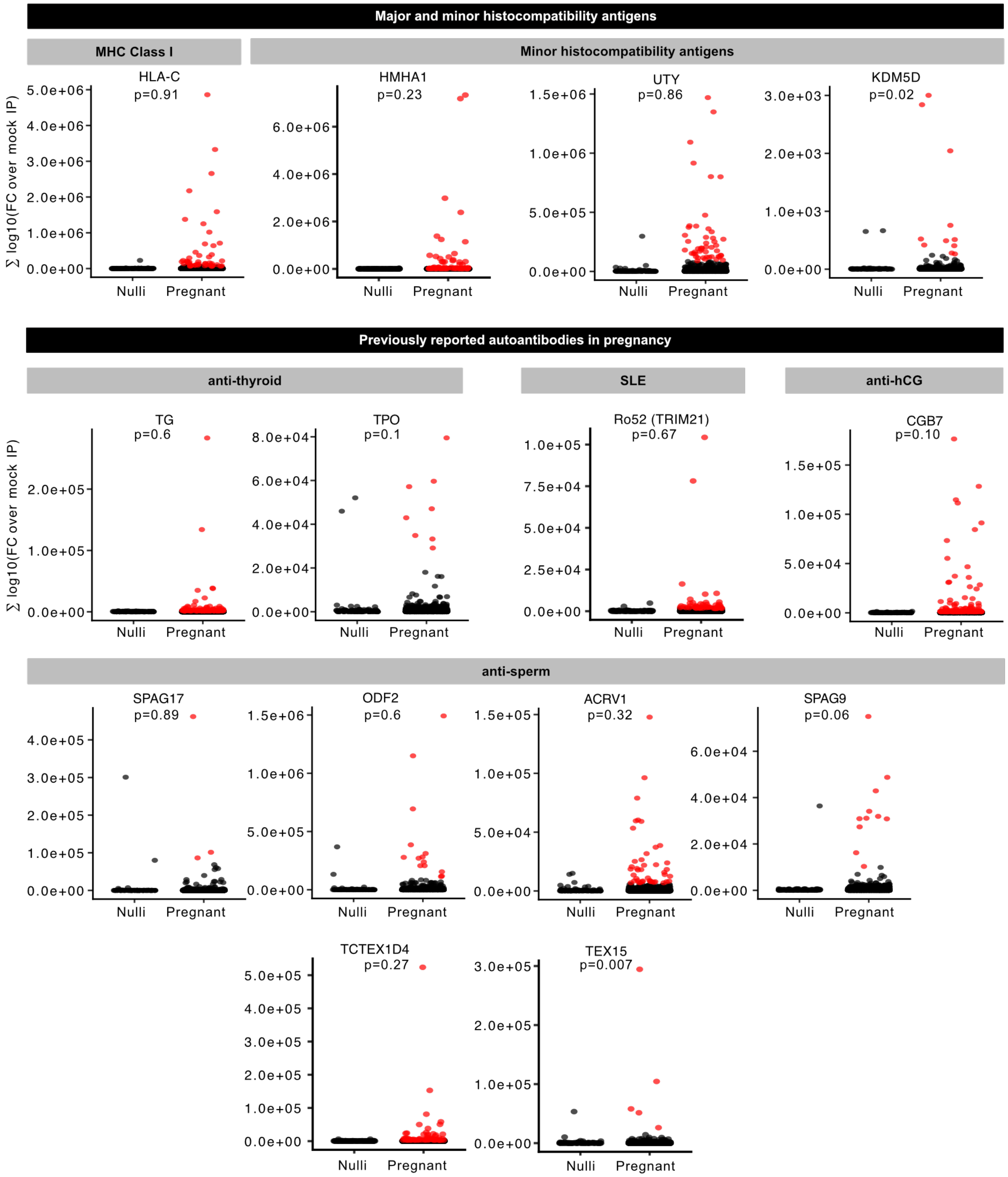


**Extended Data Figure 3**. **Autoreactivities enriched in pregnancy.** Dot plots of summed log10 fold change of mock IP within an individual for peptides belonging to proteins in indicated categories in nulligravida or pregnant sera. Red dots denote greater than 3 standard deviations from non-pregnant experiments. Individual human samples are represented by each dot, Kolmogorov-Smirnov pairwise test for significance.

#### Extended Data Figure 4

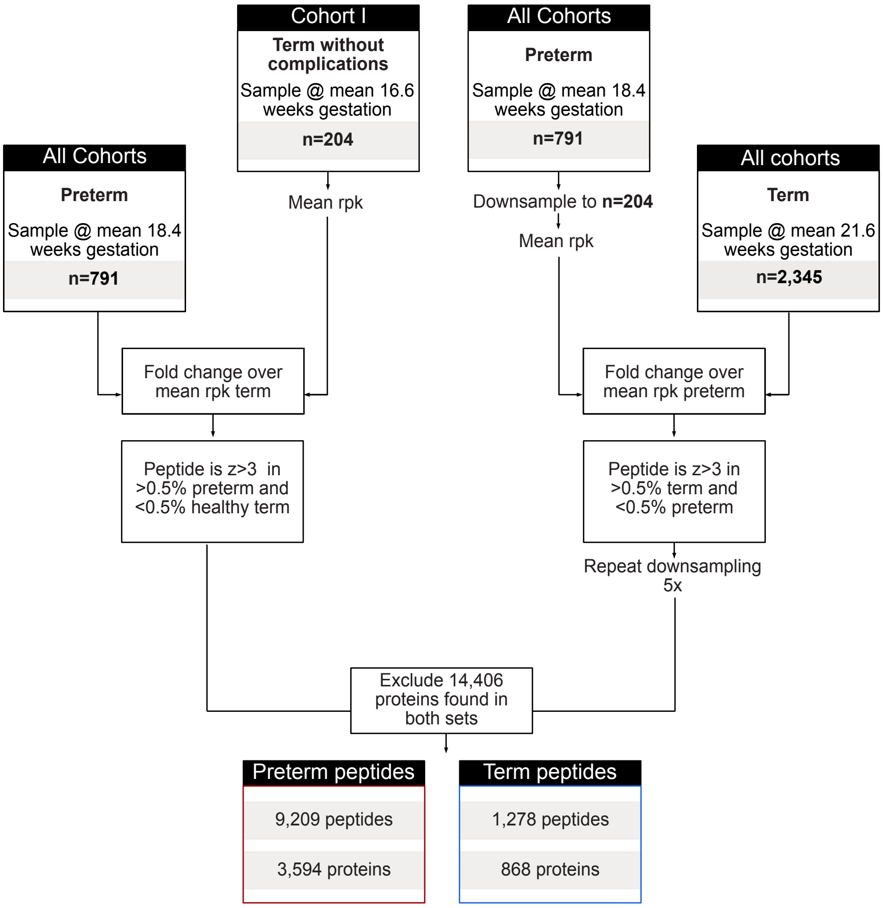


**Extended Data Figure 4. Identification of term- and preterm-specific autoreactivities by PhIP-seq**. Flow chart of hit filtering strategy from input peptide library to term and preterm specific hits.

#### Extended Data Figure 5

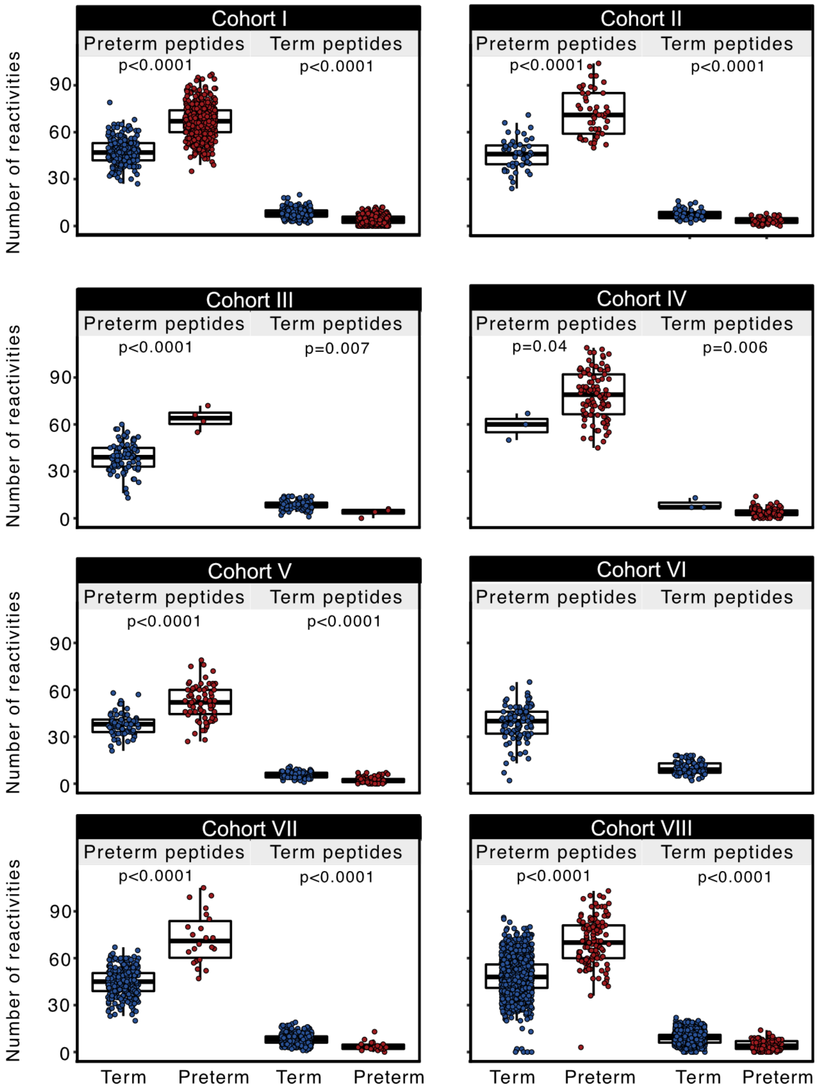


**Extended Data Figure 5. Preterm autoantibody signature across cohorts.** Number of preterm (left) and term (right) specific autoantibody reactivities in term (blue) and preterm (red) pregnancies across all cohorts. Individual human samples are represented by each dot, Kolmogorov-Smirnov pairwise test for significance.

#### Extended Data Figure 6

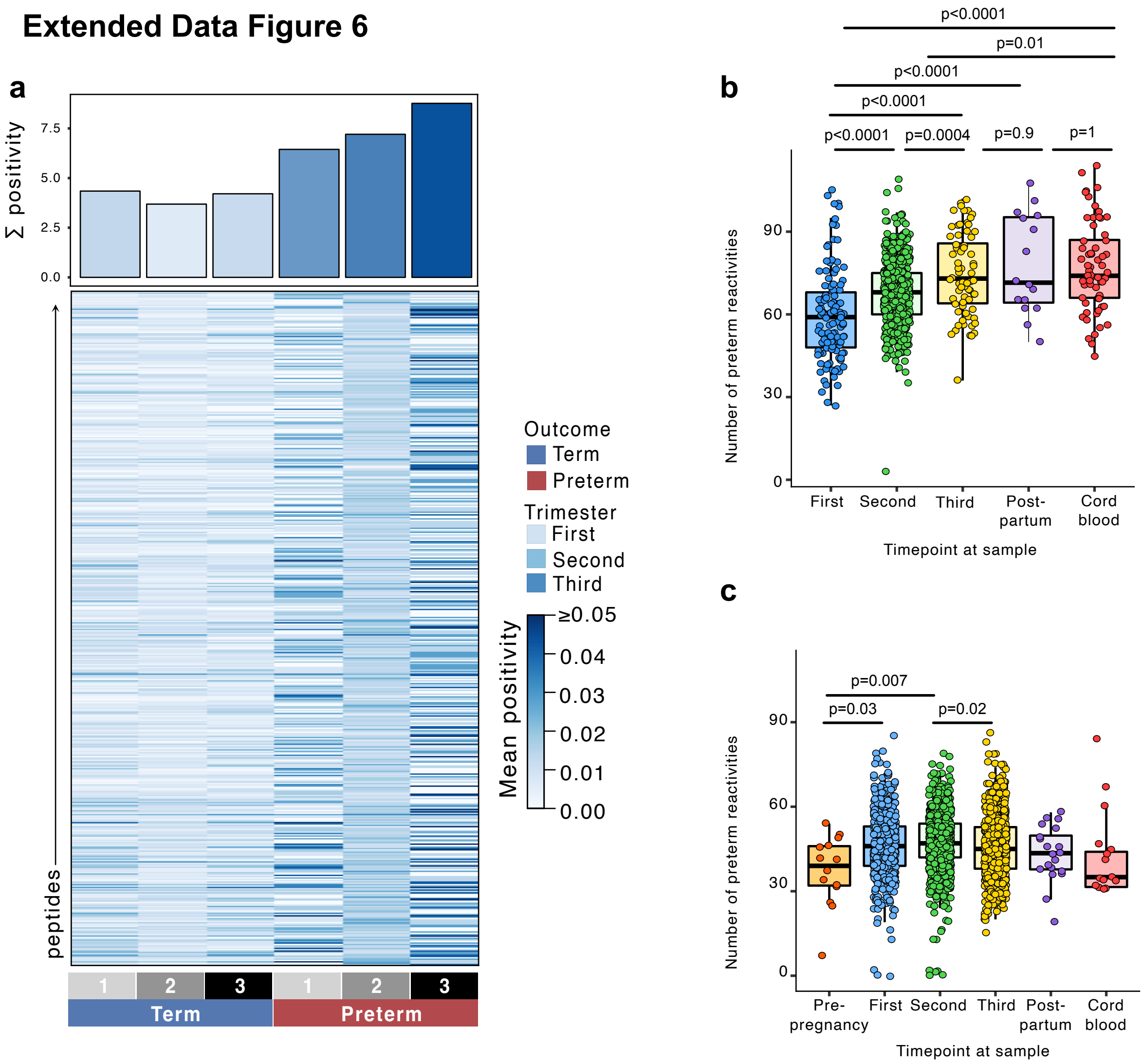


**Extended Data Figure 6. Preterm autoantibody signature across trimesters. a.** Heatmap colored by mean positivity (of individual binarized peptide hit calls) of the top 500 peptides in the preterm autoantibody signature in term or preterm pregnancies in the first (1), second (2), or third (3) trimesters. Barplot (top) indicates sum of positivity across all peptide means. Boxplot of the number of preterm reactivities per sample in **b** preterm or **c** term individuals grouped by timepoint of collection. Enrichments were identified by PhIP-seq in a-c; individual human samples are represented by each dot in b-c; ANOVA test for significance with Tukey post-hoc in b-c, with selected comparisons represented in the graph.

### Extended Data Figure 7

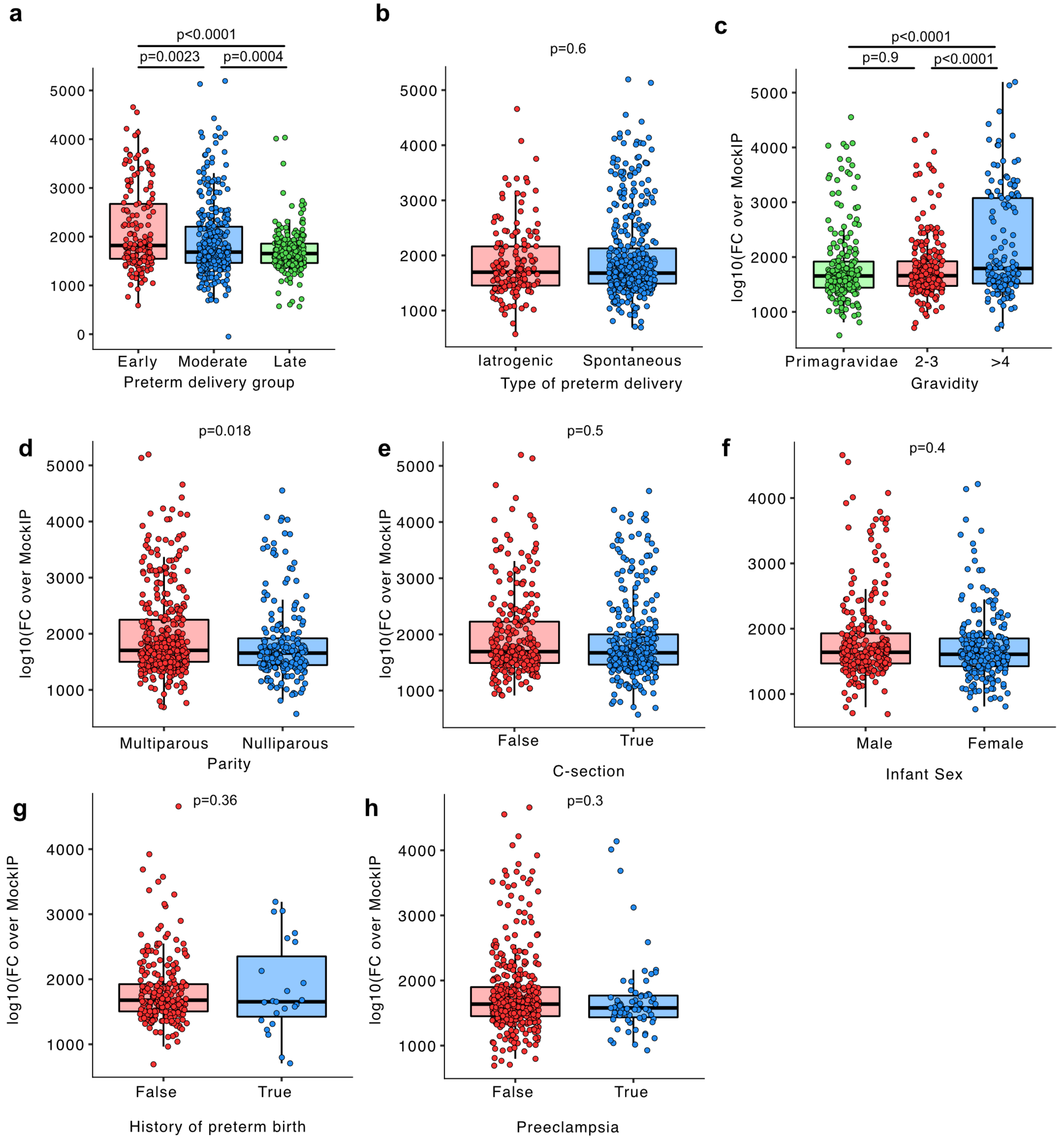


**Extended Data Figure 7. Preterm autoantibody signature with respect to clinical variables.** Summed log10 fold change over mock IP of the preterm-specific autoantibody signature in preterm pregnancies stratified by **a.** early (<28 weeks), moderate (28-32 weeks), and late (33-37 weeks) delivery age groups, **b.** type of preterm delivery, **c.** gravidity categories, **d.** parity, **e.** delivery mode, **f.** male or female infant sex, **g.** reported history of preterm birth, **ih.** preeclampsia diagnosis. One-way ANOVA with Tukey HSD in a, c, Kolmogorov-Smirnov pairwise test in b,d-h for significance. Individual human samples are represented by each dot. Participants for which clinical variables were not available were excluded from the analysis.

#### Extended Data Figure 8

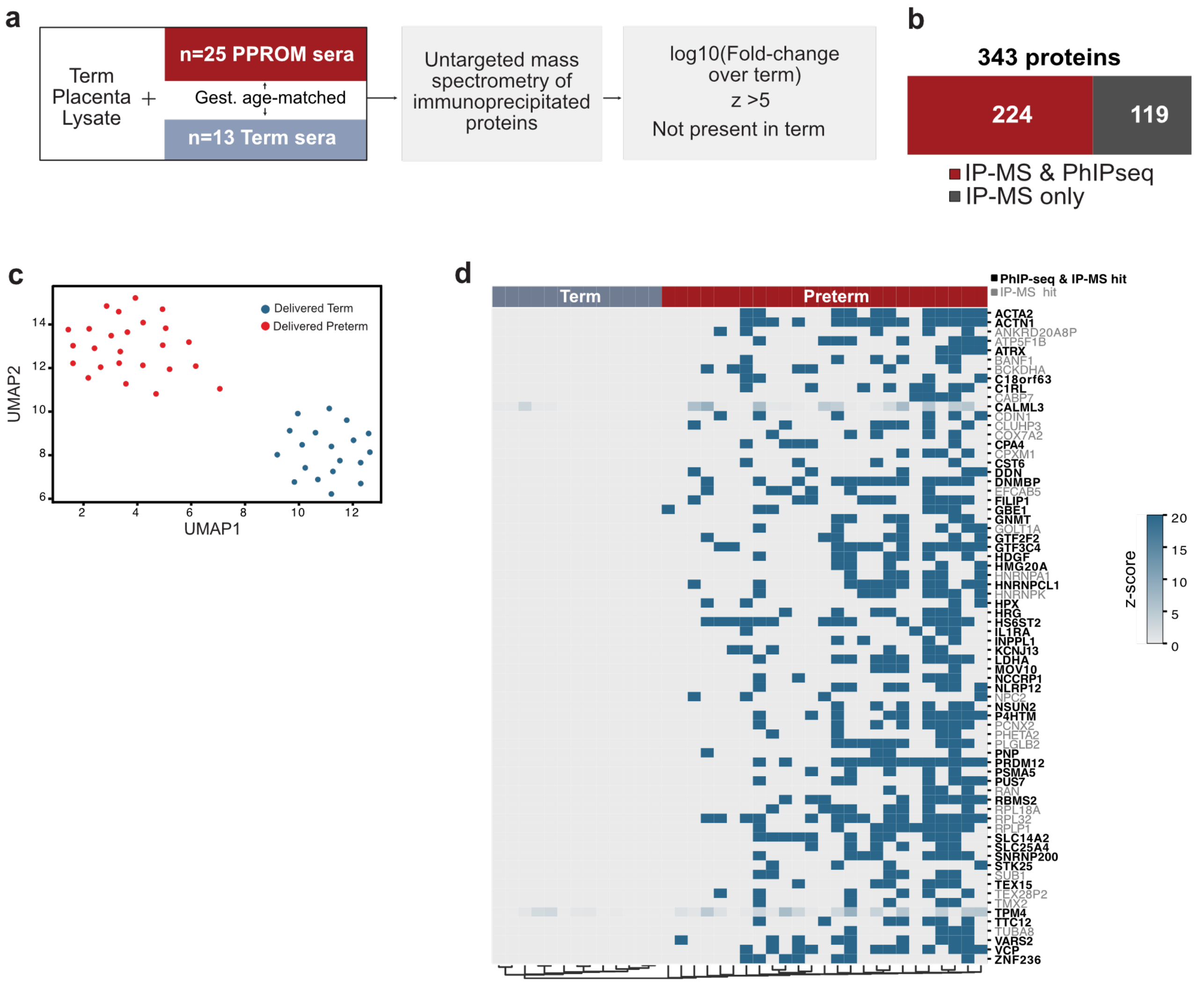


**Extended Data Figure 8. Validation of preterm-specific autoantibody signature. a.** Schematic of the placental IP-MS experiment and hit filtration approach (left) **b.** Barplot of preterm-specific autoreactivities in IP-MS analysis that overlap with those identified by PhIP-seq for Cohort II. **c.** Unsupervised UMAP analysis of placental IP-MS protein abundances in Cohort II, colored by subsequent term (blue) or preterm (red) delivery. **e.** Heatmap of hierarchically clustered z-scores from top 70 preterm-specific reactivities by placental IP-mass spec. Bolded protein names indicate that the hit was also identified by PhIP-seq, while gray protein names indicate the hit was identified by IP-MS only.

#### Extended Data Figure 9

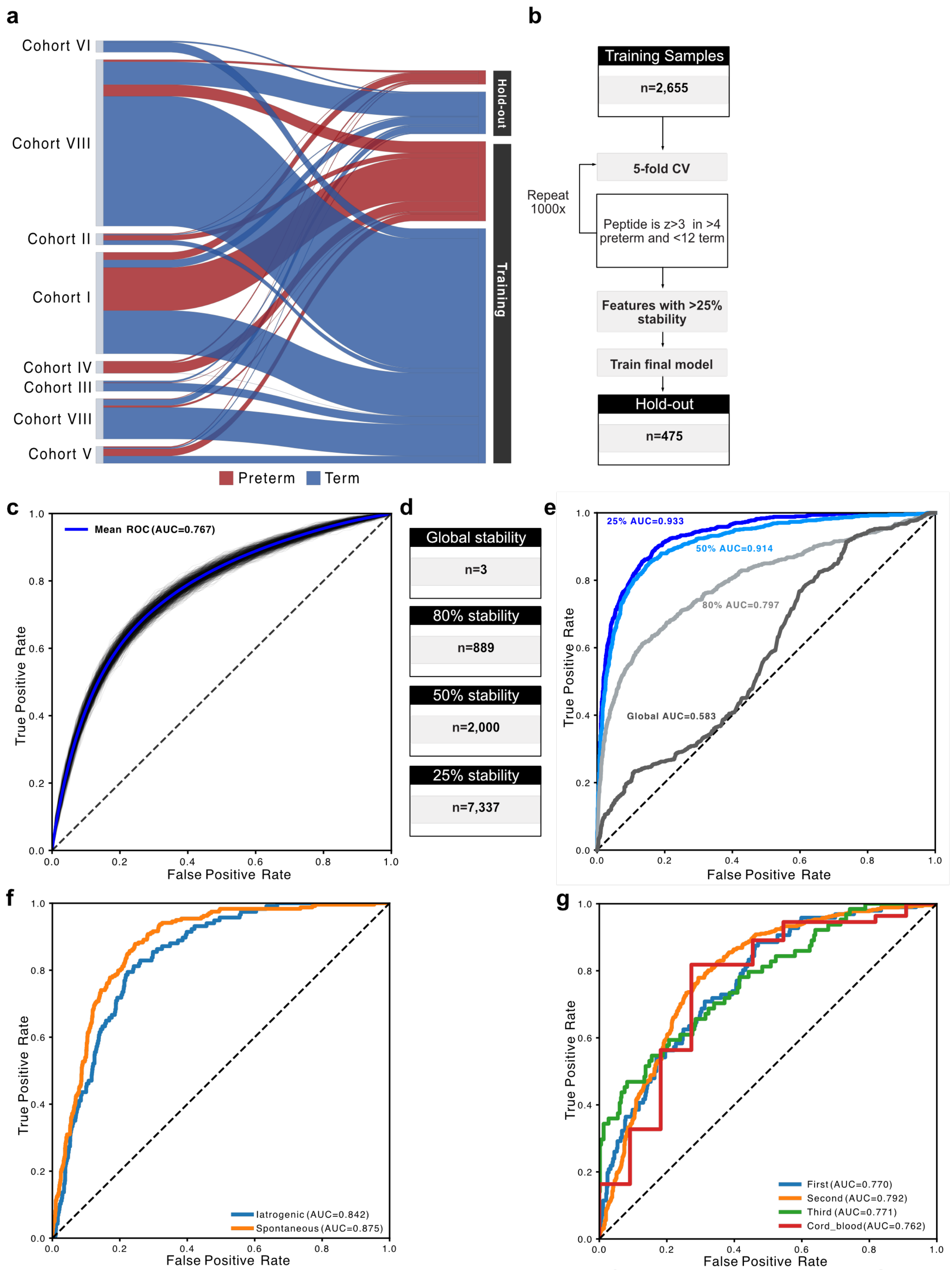


**Extended Data Figure 9. Machine learning predictor for preterm delivery. a.** Schematic of samples designated for training or testing from each cohort with term or preterm outcomes. **b.** Schematic of samples used for feature selection and final model training. **c**. Receiver operator curve analysis of feature 1000 iterations of five-fold feature selection. **d**. Schematic representing the percent of features that appeared in 1000 iterations. Receiver operator curve analysis of out-of-fold prediction across **e.** feature stability metrics, **f.** types of preterm delivery, and **g.** timepoint category at sampling for the training set.

#### Extended Data Figure 10

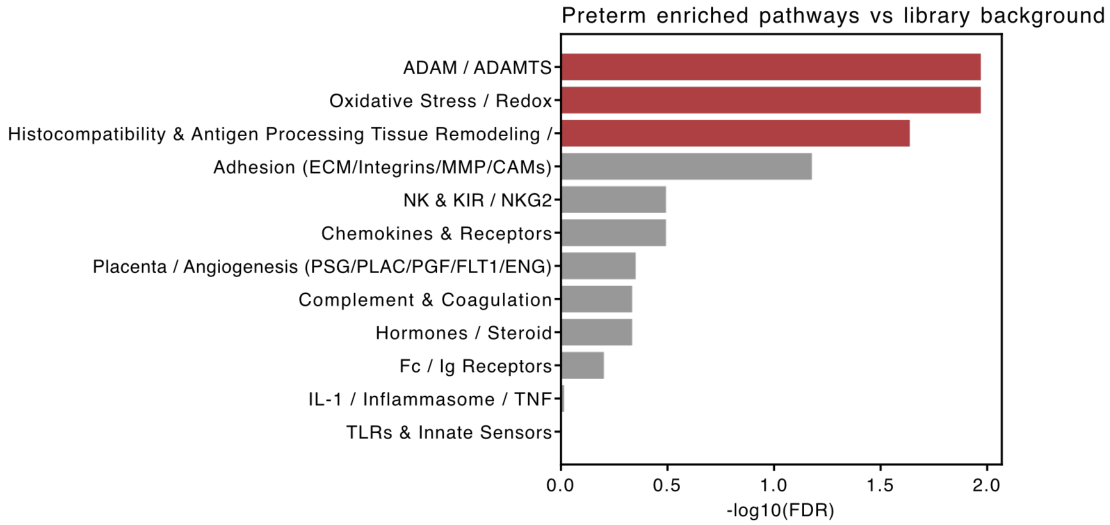


**Extended Data Figure 10. Gene-set enrichment analysis of preterm-specific autoreactivities**. Barplot of -log10 adjusted p-value of preterm-specific autoreactivities within pathway-specific gene lists. Red bars indicate significant enrichment relative to library background.

#### Extended Data Figure 11

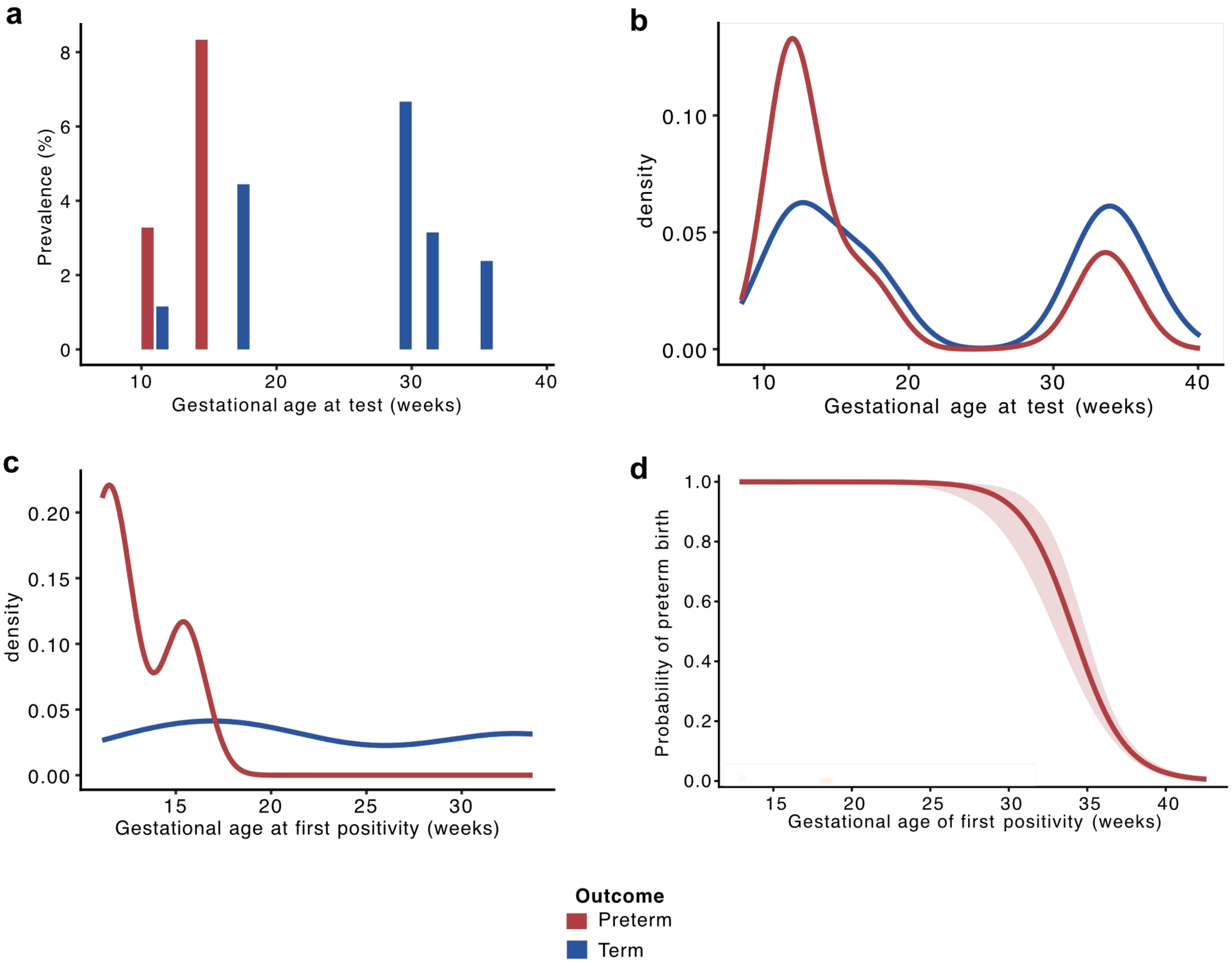


**Extended Data Figure 11. Prevalence of IL-1RA autoantibody positivity**. **a.** Prevalence of anti-IL1RA positive pregnancies binned by gestational age at sampling (2-week bins). Bars show the proportion of positive samples within each bin. Only bins with ≥2 total samples were included. Density of anti-IL1RA positive pregnancies with respect to **b.** gestational age at testing or **c.** gestational age at first detectable IL-1RA autoantibody positivity in subjects who became positive at least once. For each individual, positivity was defined as the earliest gestational age at which anti-IL1RA was detected. **d.** Logistic regression predicted probability of preterm delivery across gestational age at first detectable positivity of anti-IL1RA with never-positive individuals assigned their gestational age at delivery. The shaded region represents the 95% confidence interval. Pregnancies were considered to be anti-IL1RA positive if the MFI over BSA exceeded three standard deviations from the mean in term pregnancies. Preterm and term outcomes are represented in a-c.

**Extended Data Figure 12**

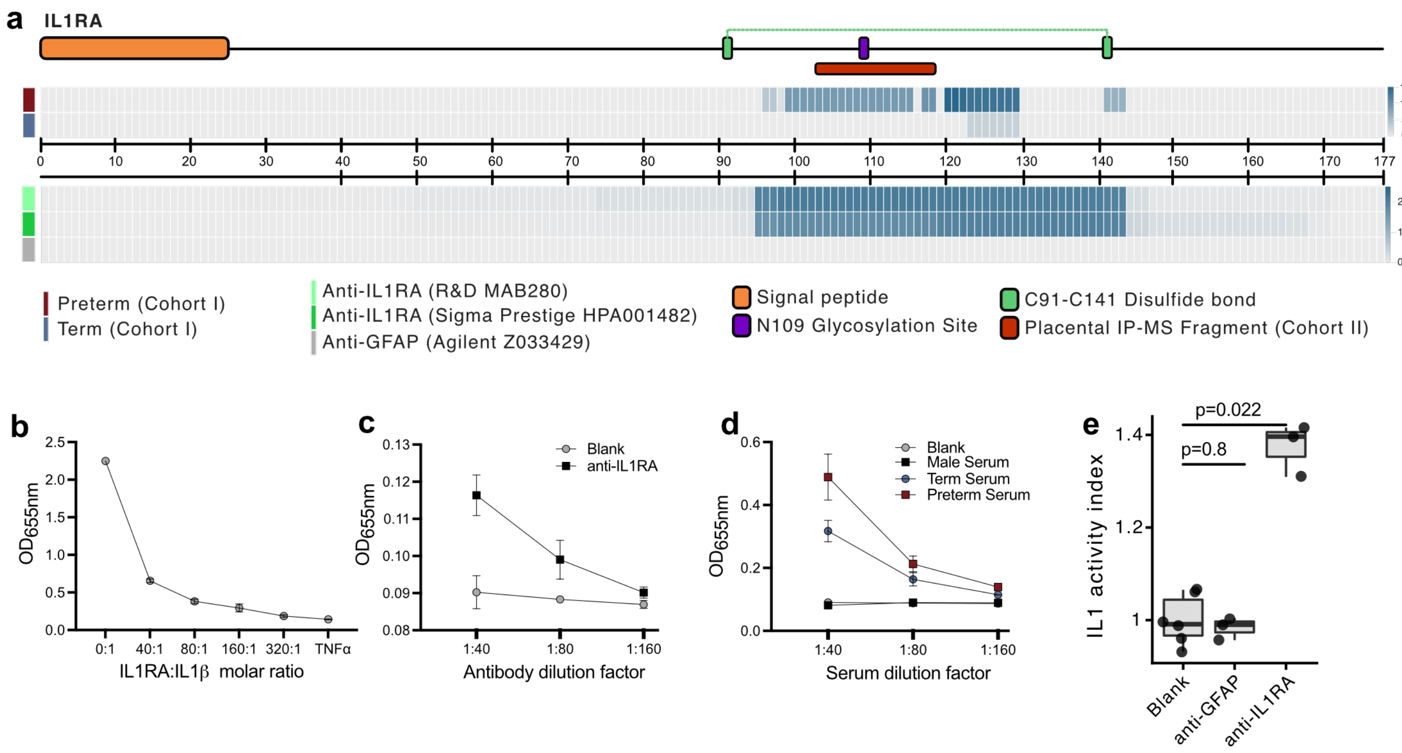


**Extended Data Figure 12. Neutralizing anti-IL1RA antibodies dysregulate IL1**. **a.** Domain structure of IL1RA with placental IP-MS fragment enriched in preterm pregnancies in Cohort II (red; top). Heatmap of mean corresponding positional enrichment for each amino acid by PhIP-seq in term and preterm pregnancies in Cohort I (middle) or in IPs with commercial anti-IL1RA or off-target anti-GFAP antibodies. SEAP activity in HEKBlue IL1 reporter cell line as measured by the conversion of a chromogenic substrate at $OD_{655}$ in titration experiments of **b.** decreasing IL1RA **c**. anti-IL1RA antibody, or **d.** sera from a male donor or term or preterm pregnancy. **e.** Quantification of secreted embryonic alkaline phosphatase (SEAP) produced by HEK-Blue IL-1β reporter cells incubated with human IL-1β and IL-1RA in the presence of PBS, off target anti-GFAP, or anti-IL1RA polyclonal antibody. Statistical significance calculated by one-way ANOVA with Tukey post-hoc in e.

#### Extended Data Figure 13

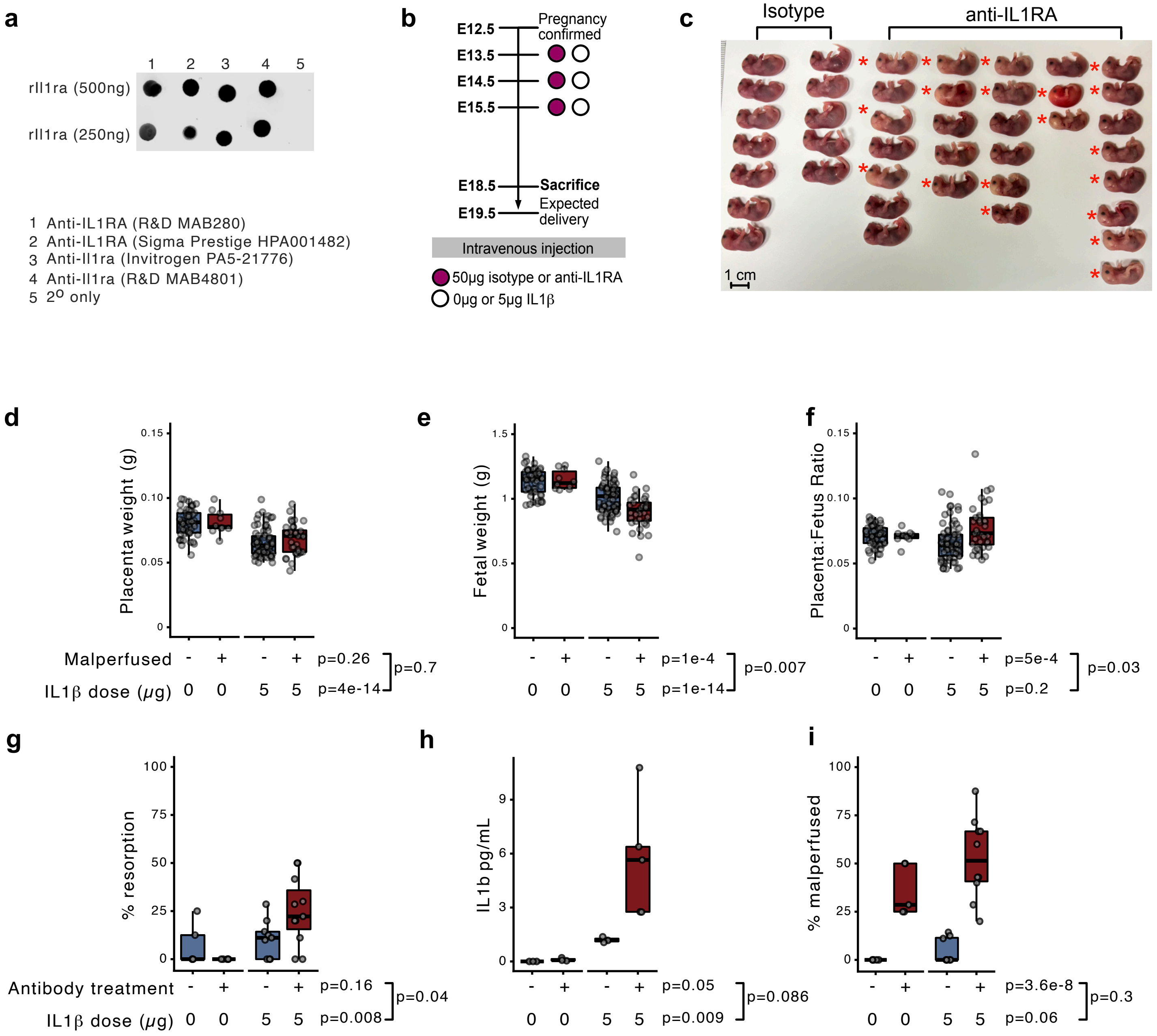


**Extended Data Figure 13. Neutralizing anti-IL1RA antibodies dysregulate IL1 *in vivo*.**
**a.** Dot blot of recombinant mouse 500ng (top) or 250ng (bottom) Il1ra probed with anti-human IL1RA antibodies (lanes 1-2), or anti-mouse Il1ra (lanes 3-4), or secondary only control (lane 5). **b.** Schematic of injections in timed-pregnant C57Bl/6 mice. Isotype or anti-IL1RA commercial antibody was injected daily on embryonic days (E) 13.5-15.5 with either 0 or 5μg of IL1; mice were sacrificed on E18.5. **c**. Representative images of fetuses from isotype or anti-IL1RA injected mice. Each column represents pups from one dam, asterisk indicated observation of gross vascular defects. Quantification of **d.** placenta weights, **e.** fetal weights, and **f.** placenta:fetus weight ratio in malperfused or normal pups relative to IL1 dose. Quantification of **g.** the percent resorption, **h.** maternal serum Il1b levels (pg/mL) and **i.** percent malperfusion in pregnancies treated with anti-IL1RA (+) or isotype (-) and relative to IL1 dose. Two-way ANOVA test for significance and interaction in d-i.

#### Extended Data Figure 14

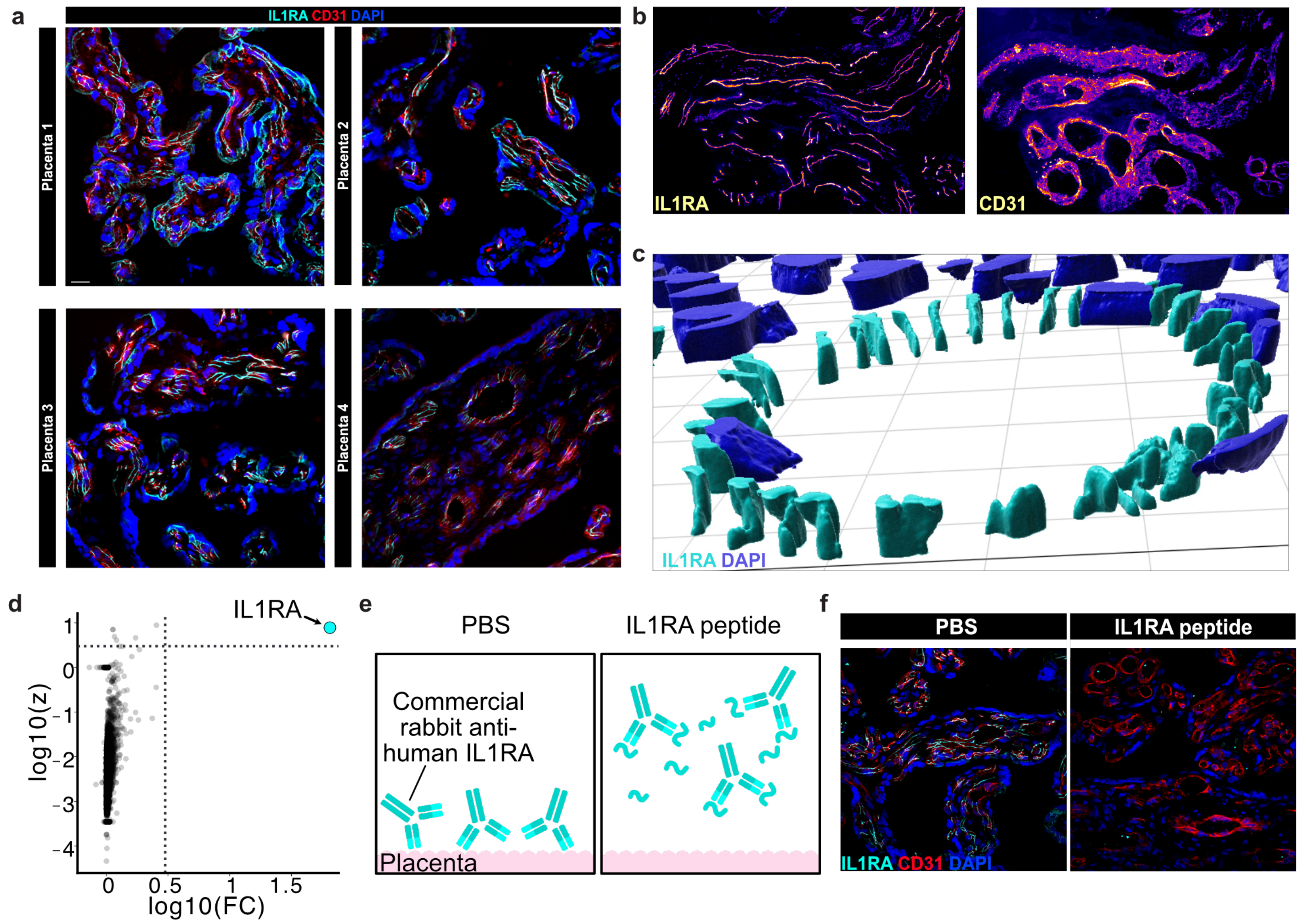


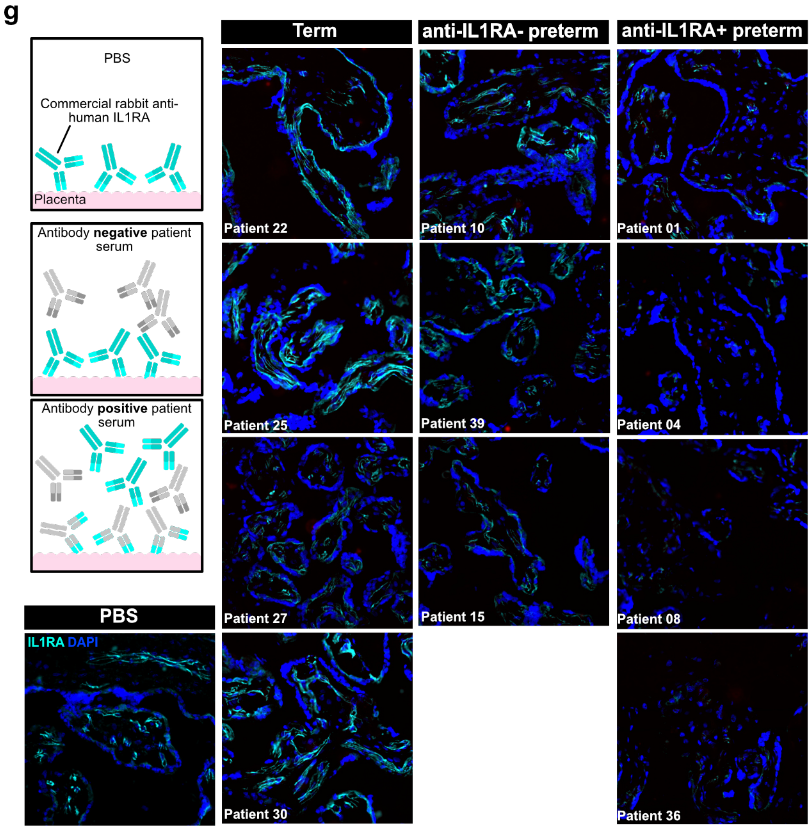


**Extended Data Figure 14. IL1RA localizes to placental vessels**. **a.** Spinning disk confocal images of indirect immunofluorescence of IL1RA (cyan) and CD31 (red) in n=4 term human placentas. Composite images with DAPI counterstain. Scale bar indicates 10μm. **b**. Single channel emission of representative placenta stained with anti-IL1RA (left) or CD31 (right). **c.** Space-filling model of a three-dimensional reconstruction of a placenta vessel immunofluorescence probed with anti-IL1RA (cyan) and counterstained with DAPI (blue). **d.** Volcano plot of PhIP-seq enrichments utilizing commercial anti-IL1RA antibody represented by log10(z-score above mock IP) and log10(fold change over mock IP). Each dot represents one protein in the PhIP-seq library. **e**. Schematic of peptide blocking experiment. **f**. Indirect immunofluorescence of IL1RA and CD31 in experiments with (right) and without (left) a blocking peptide. Representative images of three experiments. **g**. (inset) Schematic of blocking experiment with patient sera and counterstaining with commercial anti-IL1RA antibody. Indirect immunofluorescence of IL1RA (cyan) in placental sections pre-blocked with anti-IL1RA negative term serum, anti-IL1RA positive preterm serum, or PBS. Each slide was blocked with serum from a different subject. Scale bar represents 50μm. Representative images of five experiments.
